# Pontine Lesions Impair Visually Guided Eye and Hand Movements

**DOI:** 10.1101/2020.05.05.20091371

**Authors:** Friedemann Bunjes, Peter Thier

## Abstract

Although animal research and some rare human case reports suggest that lesions of the dorsal pons yield saccadic and smooth pursuit eye movement deficits, little is known about the functional topology of the human pontine nuclei (PN) and whether limb movements are similarly affected as eye movements. Saccadic as well as SP eye and pointing movements were measured in six patients with lesions in the PN region. Five patients of the six exhibited dysmetric saccades, whilst smooth pursuit gain was reduced in four. Pontine lesions also alter the relationship between amplitude, velocity, and velocity skewness of saccadic eye movements. Limb movement trajectories were more curved in four patients. The results suggest that the lesions impair a general calibration mechanism that uses the parallel fiber-Purkinjecell synapse in the cerebellar cortex to adjust the timing of muscle innervation in visually guided oculomotor as well as limb movement tasks.

## Introduction

The cerebellum is necessary for well-coordinated and smooth movements of the eyes and limbs. The largest single input to the human cerebellum arises from the pontine nuclei (PN). Axons of pontine cells terminate in the cerebellar cortex as mossy fibres. PN and the Nucleus Reticularis Tegmenti Pontis (NRTP) are a major pathway relaying visual information from the cerebral cortex (Glickstein et al., 1994) and the superior colliculus (Harting JK, 1977) to the cerebellum. Among other cerebellar targets, the axons of pontine cells project to lobules VI and VII of the oculomotor vermis (OMV, Yamada J, Noda H., 1987) and the contralateral dorsal and ventral paraflocculus (Brodal P et al. 1991). Other PN projections end in the cerebellar nuclei fastigii and interpositus (Mihailoff, 1993; Shinoda et al., 1992). Neurons in the dorsolateral PN (DLPN) and NRTP of monkeys are active prior to smooth pursuit eye movemenents (Mustari et al., 1988; Ono et al., 2005; Ono et al., 2004; Suzuki et al., 1990; Thier et al., 1988) and saccades (Dicke et al., 2004). Pontine lesions in monkeys cause smooth pursuit deficits and dysmetric saccades (May et. al., 1988).

Three studies so far concentrated on the oculomotor consequences of human PN lesions. Thier et al. (Thier et al., 1991) reported on a patient showing a selective ipsiversive SP deficit and a mild saccadic hypometria. Gaymard and co-workers (Gaymard et al., 1993) found in four patients a reduced SP gain especially during ipsiversive eye movements. No saccadic deficits could be detected but only the latency and maximum velocity were analysed. Deleu et al. (Deleu et al., 1997) observed ipsiversive SP as well as saccadic deficits in patients with pontine lesions. However, the fact that the paramedian pontine reticular formation (PPRF) containing premotor oculomotor neurons was also involved in these lesions renders it impossible to make only the PN-lesion responsible for the oculomotor deficits. The publication of Schmahmann and co-workers (Schmahmann et al., 2004a) focuses on limb movements but eye movement deficits of the tested 25 PN-patients are mentioned. The authors observed normal eye movements (14 patients), gaze-directed-nystagmus (3 patients), hypo- and hypermetric saccades (contra- and ipsiversive, 7 patients) and SP-deficits (6 patients).

The role of the PN in the generation and control of limb movements has so far not been investigated as intensely as for eye movements. One electrophysiological study (Matsunami, 1987) found 63 neurons in the monkey PN (including the NRTP) that discharged in relation to wrist movements mainly during a time interval of 200 ms before to 100 ms after movement onset. Tziridis and co-workers (Tziridis et al., 2009; Tziridis et al., 2012) found a distinct cluster of neurons in the rostrodorsal PN which was activated by the preparation and the execution of hand reaches, close to but distinct from the region in which saccade-related neurons were observed. Their findings suggest a distinct precerebellar, pontine visuomotor channel for hand reaches that is anatomically and functionally separated from the one serving eye movements.

A functional study respecting the human PN has been published by Schmahmann et al. (Schmahmann et al., 2004a) in which a topographical correlation between qualitative clinical findings on motor deficits and pontine lesions of 25 patients was carried out. The lesions caused dysmetria and coordination deficits. The authors suggest a medio-ventral part of the rostral and medial pontine levels to be involved in hand movements and overlapping, but slightly more ventrally located areas for arm movements. They demonstrate a clear spacial segregation between facial and speech-related regions and those correlated with limb movements.

Although the importance of the PN for the generation and control of SP and saccadic eye movements in nonhuman primates has been definitely shown it is still unclear, if the situation is similar concerning the human PN. In this study we quantitatively demonstrate that PN lesions in humans cause saccadic and SP deficits as well as an impairment of limb movements.

## Methods

### Patients

All six PN-patients A to F showed well defined lesions in the area of the pontine nuclei. Some corticospinal as well as pontocerebellar fibers were also affected.

### PN-Patient A

Patient A (m, 64 years old) had a paramedian left-sided pons-infarct (Figure 1). The lesion initially caused dysarthria and a disturbance of fine motor skills on the right side of the body. A positive Babinski test indicated a lesion of the pyramidal tract. The experimental investigations took place 16 days after the infarct, by wich time the patient had recovered from dysarthria.

**Figure 1.**
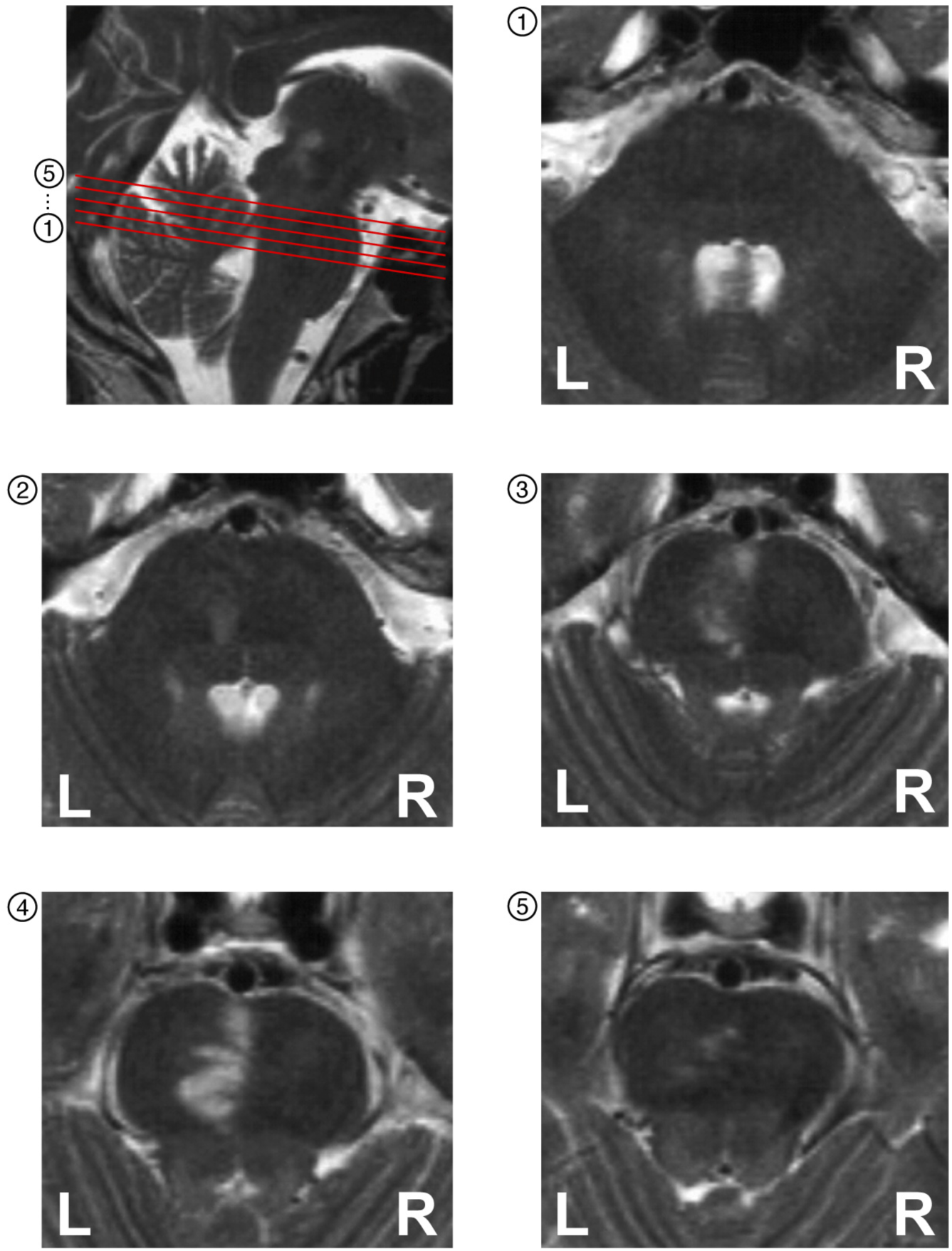
Localization of patient A’s lesion. The MRI scans of of patient A show that the lesion is located left-sided near the midline of the rostral pons.

### PN-Patient B

Patient B (m, 67 years old) suffered from gait uncertainty, awkwardness on the right body side, a spontaneous nystagmus to the right, a left-sided hemiataxia and a mild right-sided hemiparesis caused by a left-sided pons lesion (Figure 2). No pyramidal tract signs were found in this patient. Measurements were executed 16 days after the insult. Dysarthria, vertigo, paraparesis and gait uncertainty were still present.

**Figure 2.**
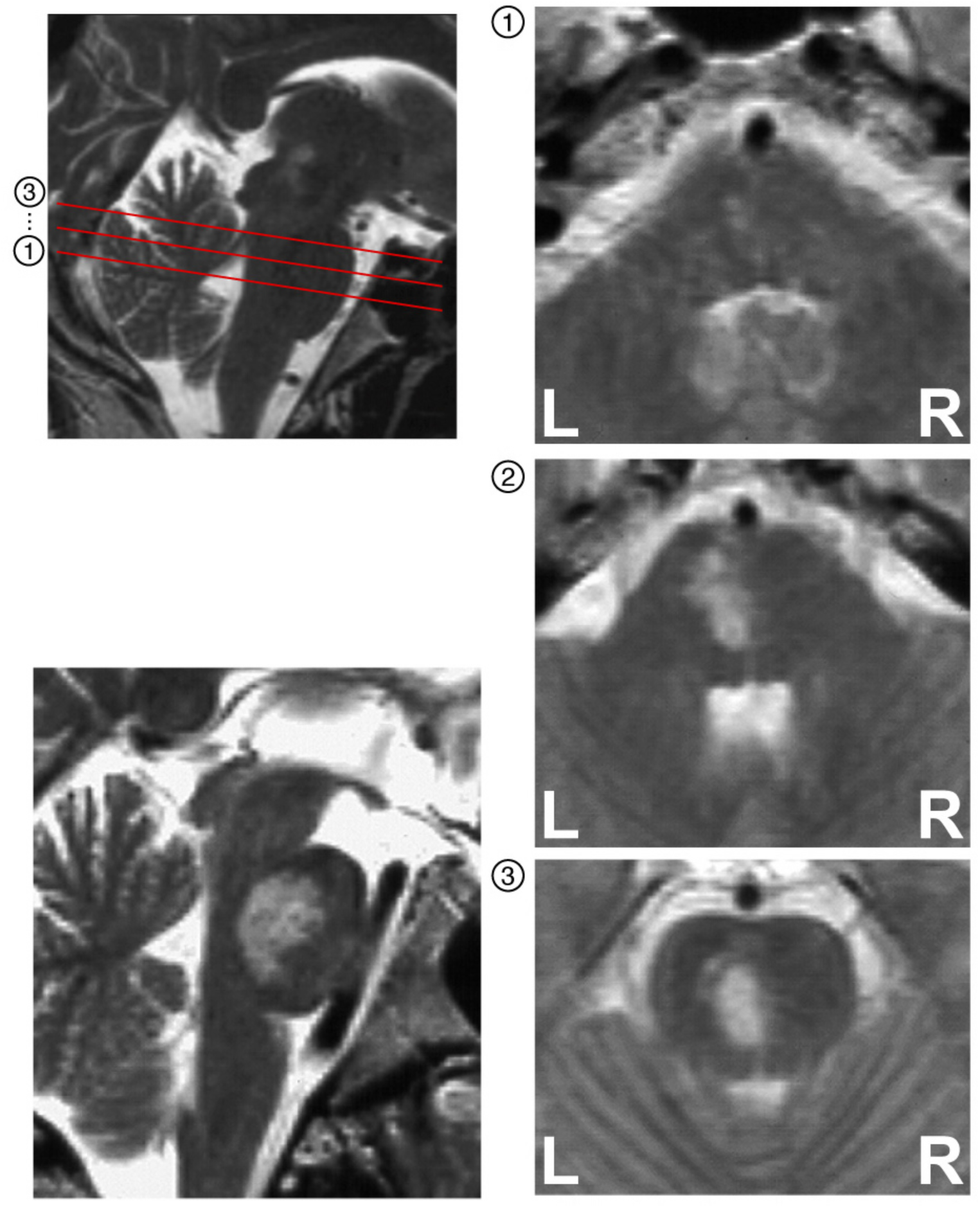
Localization of the lesion of patient B. MRI scans of the pons of patient B. Lower left image: Sagittal section showing the vertical extent of the lesion. Right images: The lesion is located on the left side of the central pons.

### PN-Patient C

Patient C (f, 42 years old) initially suffered from hemihypaesthesia, hemiparesis, disturbance of fine motor skills, facial paresis and dysarthria. These symptoms were caused by two pontine infarcts, first on the left, then on the right at an interval of 15 days (Figure 3). No pyramidal tract signs were found at this female patient either. Measurements were performed 8 days after the second infarct. At this time only the facial paresis persisted.

**Figure 3.**
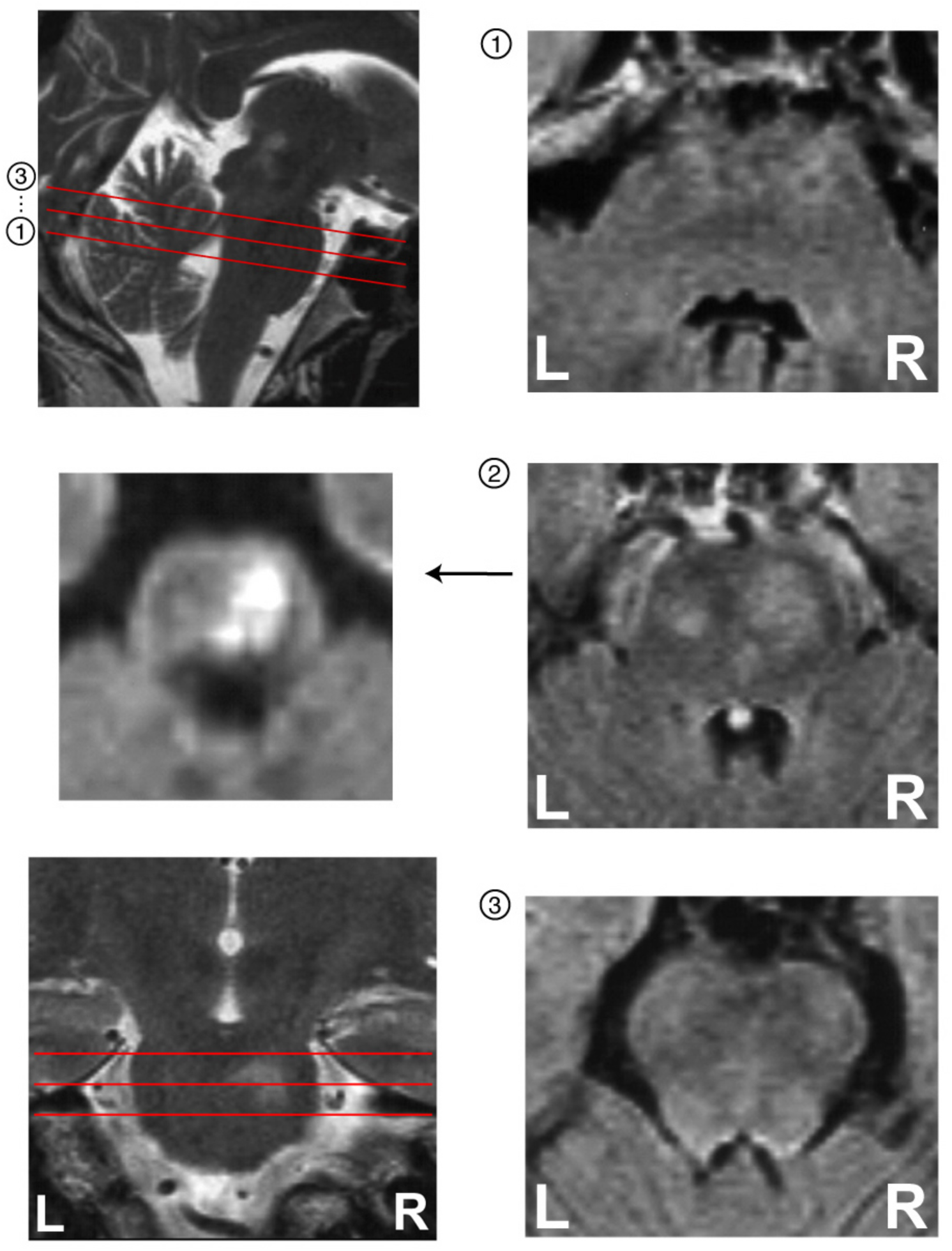
Localization of the lesion of patient C. MRI scans of the pons of patient C. Left middle image: Diffusion weighted image of section 2. Lower left image: Frontal section showing the vertical extent of the lesion. Right images: The lesion is located both sides of the central pons with a bigger extension on the right.

### PN-Patient D

Patient D (f, 76 years old) suffered from vertigo with a tendency to fall to the right and abnormal loss of weight. No pathologic reflexes were found in this patient. CT and NMR imaging revealed a tumor that was located in the left upper brainstem with a diameter of approximately 1cm (Figure 4). Eye- and limb-movement recordings took place 30 days after the first occurence of vertigo.

**Figure 4.**
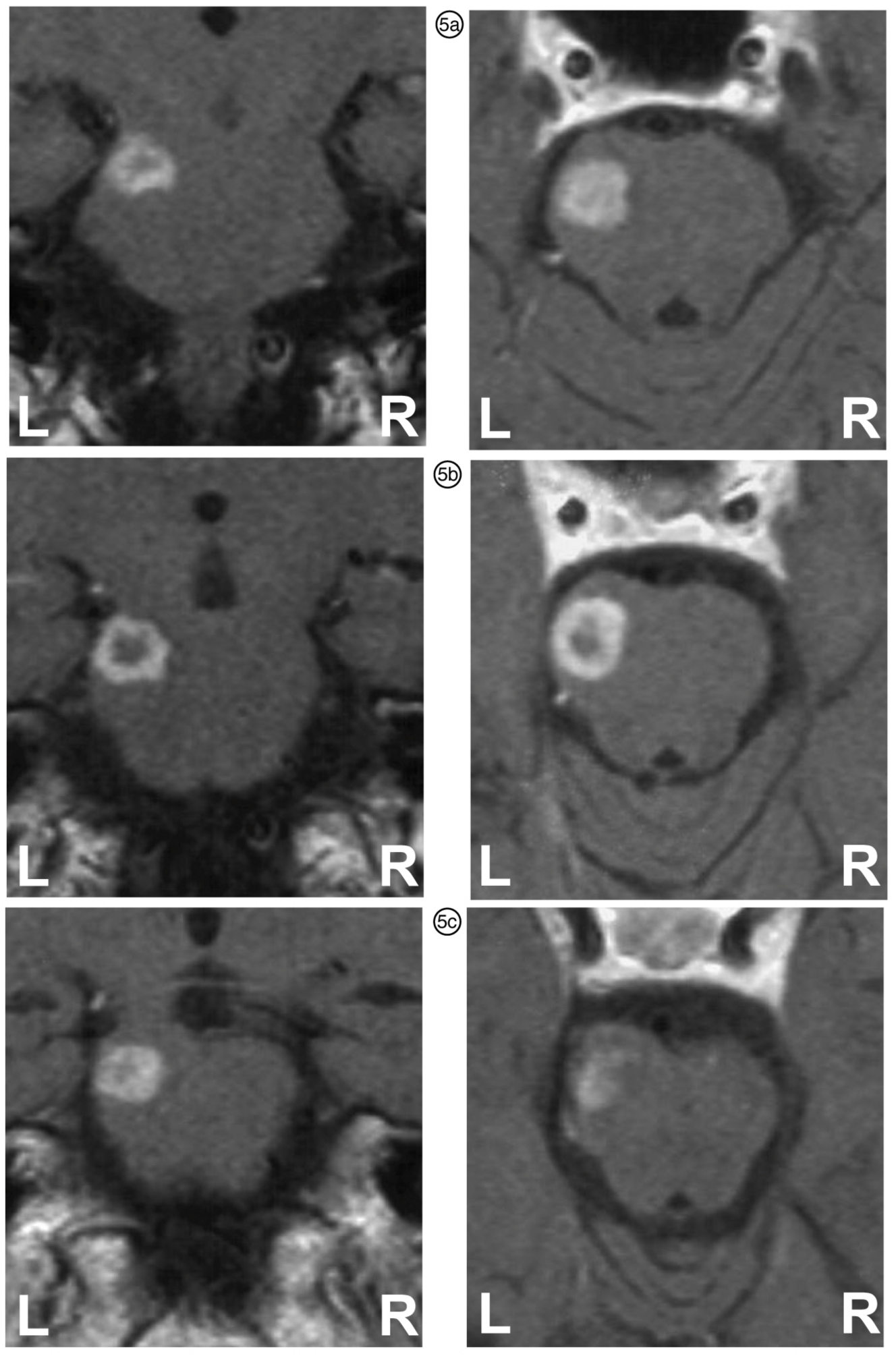
Localization of Patient D’s lesion. MRI scans of the pons of patient D. The tumor is located in the left rostral part of the pons. right: all 3 sections are located on the level of section 5 in Figure 1. Left column: frontal sections illustrating the vertical extent and the rostral position of the lesion.

### PN-Patient E

Patient E (m, 41 years old) suffered from a peripheral facial paresis with incomplete eye-closing ability on the right and a disturbance of taste perception on the right side of the tongue. Pathologic reflexes could not be detected. NMR imaging revealed a small insult in the area of the left PN (Figure 5). Patient E was studied 25 days after the initial occurence of his problems.

**Figure 5.**
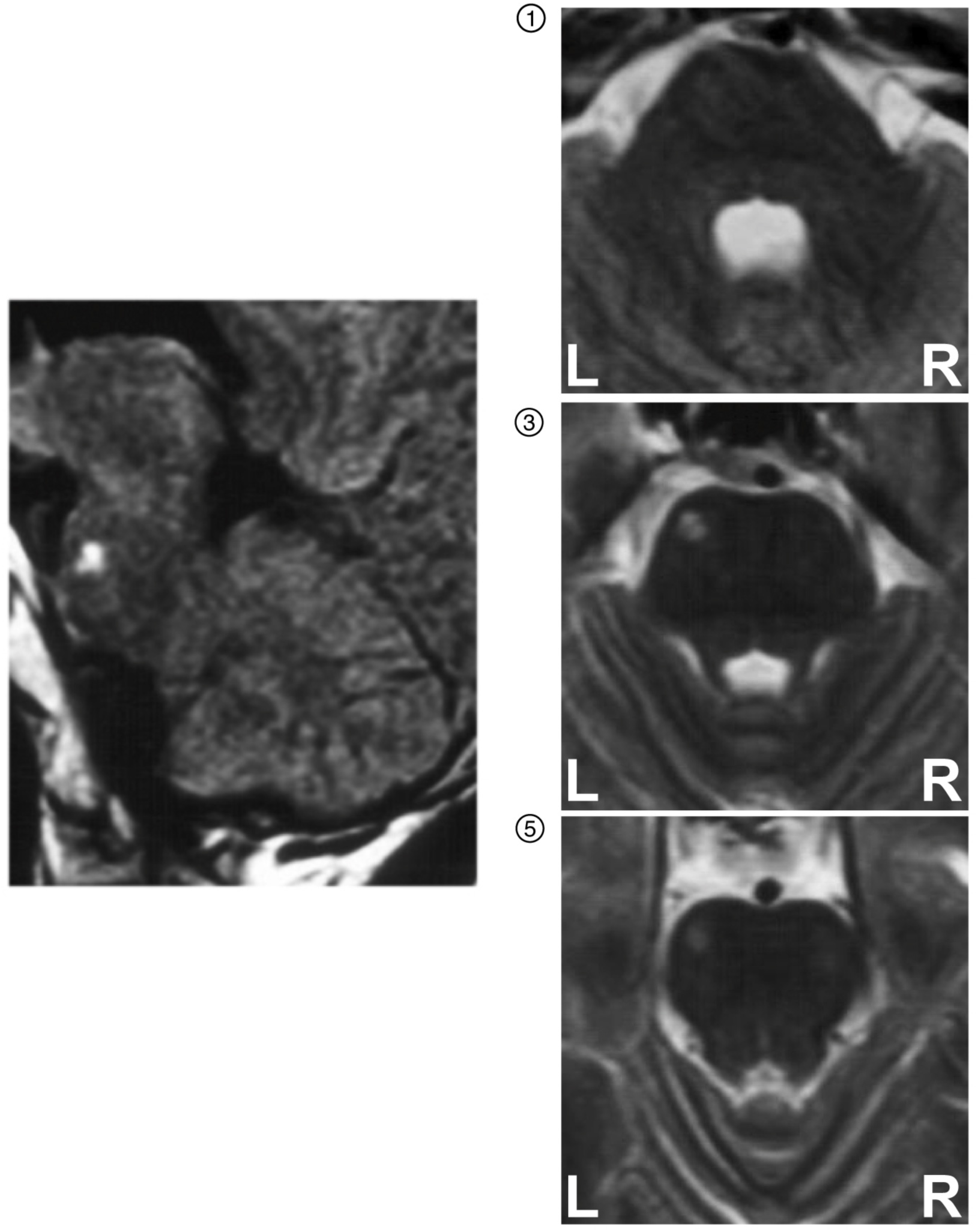
Localization of the lesion of patient E. MRI scans of the pons of patient E. The lesion is located in the left pons. right: sections correspond to layers 1, 3 and 5 in Figure 1. left: sagittal section showing the vertical extent of the relatively small lesion.

### PN-Patient F

Patient F (m, 67 years old) was affected by a brainstem insult four days before the measurements, leading to mild saccadic gaze following and saccadic hypometria to the right together with paresis of the left arm and leg, a gait distortion to the left with a tendency to fall to the left side, left-sided pointing dysmetria and bradydysdiadochokinesis. A left-sided babinski-sign was detected. The CT and MRI examination revealed an insult in the right rostral pons (Figure 6).

**Figure 6.**
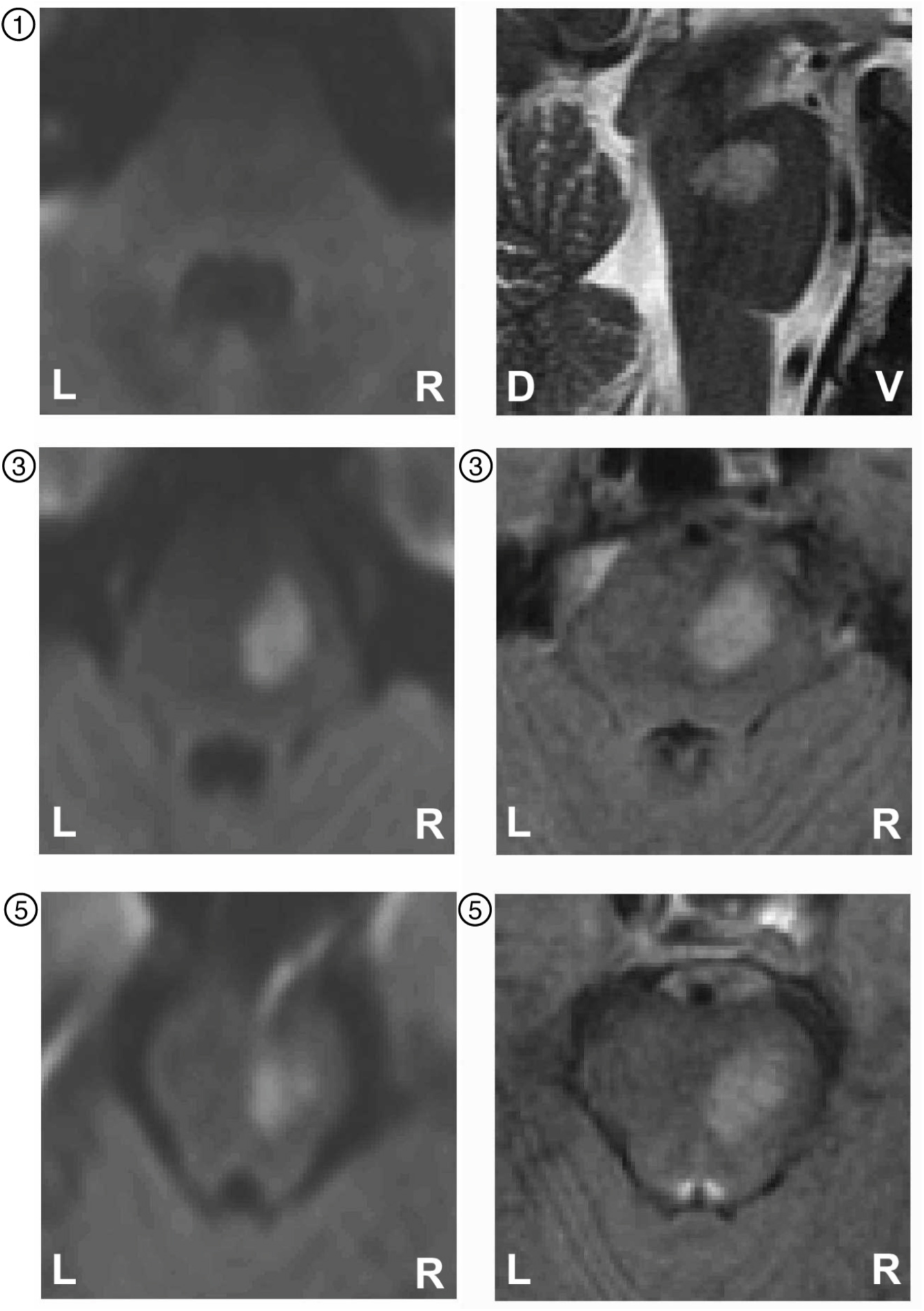
Localization of the lesion of patient F. MRI scans of the pons of patient F. The lesion is located in the right rostral pons. Left: diffusion weighted images, sections correspond to layers 1, 3 and 5 in Figure 1. Upper right: sagittal section showing the vertical extent of the lesion. Right middle and bottom: T1-weighted images of sections 3 and 5.

### Brainstem-Patient G

As a neurological control for pyramidal tract damage we also examined Patient G (f, 56 years old) who was hospitalized with a sudden neck pain, rightward vertigo and sensory deficits in the right face caused by a small ischemic lesion in the paramedian medulla oblongata due to dissection of the arteria vertebralis. (Figure 7). At the time of the investigation, 14 days after the first occurance of the symptoms, she still suffered from local headache, sickness and vertigo. The neurological investigation revealed an awkward one-foot-hopping on the right, a right-sided hypermetric finger-to-nose-and awkward knee-heel-movement.

**Figure 7.**
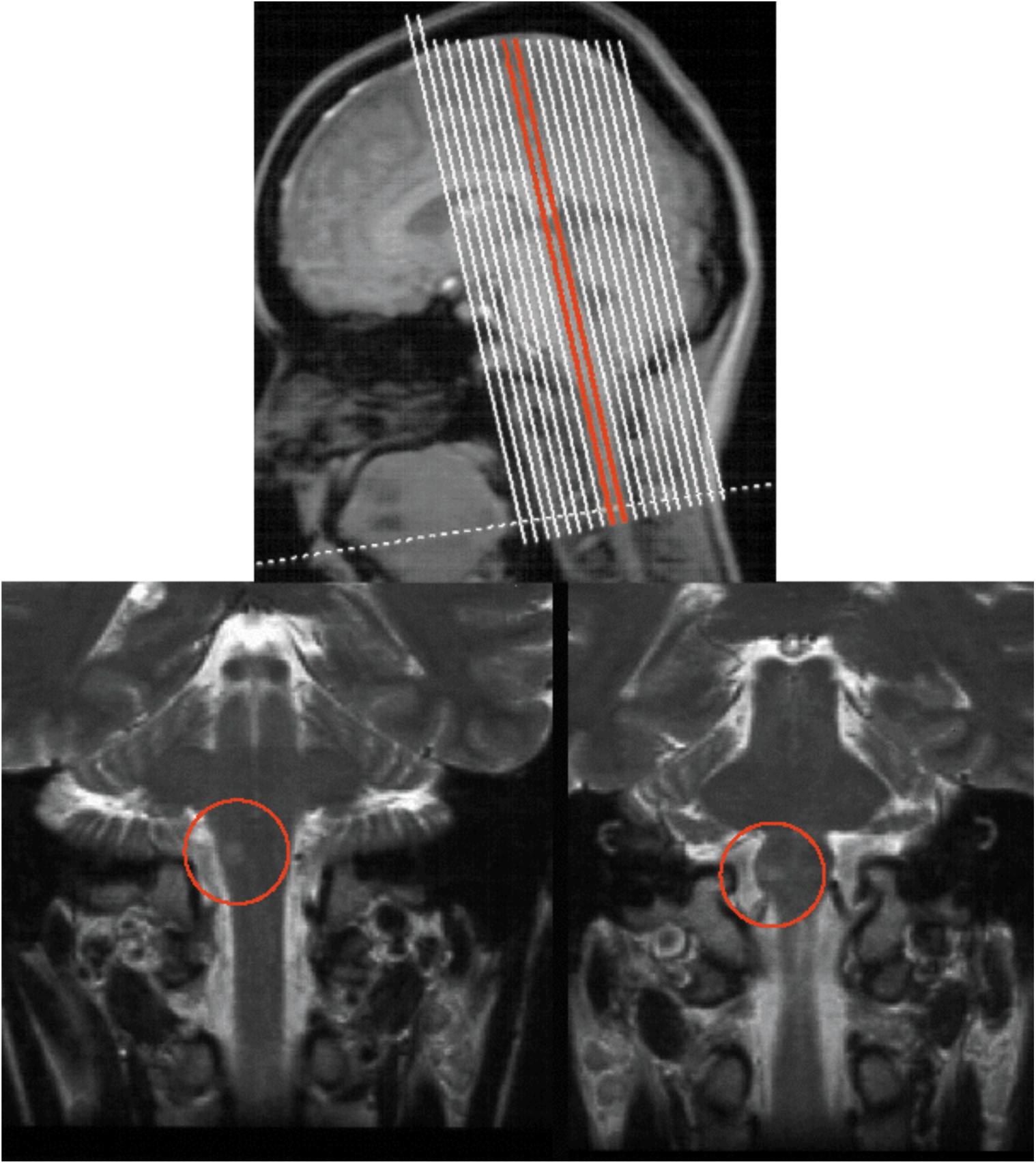
Localization of the lesion of patient G. MRI scans of the cerebellum and brainstem of patient G. Lower images: Frontal sections showing the left-sided ischemic lesion of the pyramid. Upper image: Position of these two sections.

### Control Subjects

For each patient, a group of four healthy control subjects were examined. The birthdates of the patients and the corresponding control subjects maximally differed 8 years (13 years for patient D).

Informed consent was obtained from all patients and control subjects according to the Declaration of Helsinki and to the guidelines of the local ethics committee of the medical faculty of the University of Tübingen, which had approved the study.

### Visual stimuli

All experiments were carried out in a dimly lit room. Subjects and patients were sitting on a chair in front of a computer monitor. Their head was restrained by a bite bar for movement minimization.

After a calibration sequence for the eye-movement measurement system the subjects were instructed to follow a visual target as exactly as possible. The target was a vertically oriented red bar (extension 4×12 minutes of arc, luminance 0,2 cd/m^2^)

#### Saccades

During each trial the target was kept in the center of the screen for 1500ms, and then moved to a horizontally excentric position at ±12 or ±6 degrees for 750ms. Eye position was sampled with 250Hz during 1000ms and 2250ms after appearance of the central fixation target. Excentric target positions were presented 5 to 10 times in a pseudo-random order.

#### Smooth pursuit

For the analysis of smooth pursuit eye movements, the target was moved horizontally for 35 seconds according to a sine function with a frequency of 0,5 Hz and an amplitude of 13,28 deg. Data of the first 5 seconds were rejected.

### Recording and analysis of eye movements

Eye movements were recorded using an infrared reflection system. Eye position was sampled at a rate of 250 Hz.

The records were stored on a personal computer and analysed off-line. Prior to analysis the eye recordings were filtered using a Savitzky-Golay filter (Press et al., 1992; Savitzky and Golay, 1964) with a cut-off-frequency of 27.8 Hz.

Saccades were detected using an acceleration threshold criterion. The saccade was defined as the eye position during the time interval where the absolute eye acceleration exceeded 500 deg/s^2^. Within this interval latency, duration, amplitude, maximum velocity, gain and velocity skewness were calculated. Skewness was defined as the contrast between eye acceleration time *t_a_* and deceleration time *t_d_*.

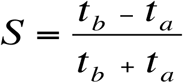

Before calculating this_€_ ratio, the eye velocity signal was resampled with interpolation at 2500 Hz to overcome the limitations of the recording sampling rate of 250 Hz. The SP gain was calculated as the ratio of the maximum eye and target velocities after saccade elimination.

### Pointing movements

During the measurement of pointing movements, the subjects were sitting in front of an array of 4 green and 1 red target LEDs that were located at eye’s height at visual angles of ±11,25 deg (red), ±22,5 deg (red) und 0 deg (green). The distance between the eyes and the central green LED was 70 cm. In the starting position, the hand was located at hip level with a horizontally oriented forearm and the elbow joint forming a right angle. One trial was performed as follows:

1. green LED on, signal tone
2. 3 seconds pause
3. green LED flickering
4. 1 second pause
5. green LED off, one of the red LEDs on, signal tone
6. measurement start
7. pointing movement towards the red target LED
8. after 3 seconds: target LED off, measurement stop, signal tone

The four target positions were chosen 5 to 10 times each in a pseudo-random order. All patients were tested with the right hand, patients B, C, D, E and G with the left as well.

### Recording and analysis of pointing movements

Hand movements were recorded using an ultrasonic 3D-movement tracker (zebris Medical GmbH, Max-Eyth-Weg 42, D-88316 Isny). A marker was mounted on the tip of the subjects’ index finger. Its three-dimensional position was sampled at a rate of 55 Hz. From the recorded data, the tangential hand velocity and, as a measure for the movement curvature, the maximum perpendicular distance between marker position and the line connecting start and end point of the movement, were calculated.

### Statistical evaluation

Differences in patient and control data were statistically evaluated using a nonparametric Wilcoxon signed-rank-test.

## Results

### Hypometric saccades

The dysmetry of the PN-patients’ saccades is illustrated in the eye movement examples in figure 8. Patients A, B, C and D showed hypometric saccades (only contraversive for patients A and C), while at patient E a greater variability and hypermetria of corrective saccades occured and patient F exhibited a mild saccadic hypermetria. Statistical analysis (Figure 9) showed significantly lower saccadic gains for both directions at patients B and D and for contraversive saccades at patients A (rightward saccades concerned) and C (leftward saccadic hypometria, contraversive relative to the bigger and latter one of the two lesions). Although patient E’s gain was not significantly different to the control data, a hypermetria of leftward corrective saccades could be observed. Patient F’s saccades showed mild but significant hypermetria. Patient G with a lesion in the pyramid exhibited normal saccadic gains. The size of the corrective saccades relative to the initial saccade size (amplitude ratio) was significantly higher for PN patients A to E in both directions (Figure 10). While the corrective saccades of the controls had a mean amplitude that was approximately 10% of the initial saccade amplitude, these PN patients exhibited a ratio of 15-25%. Amplitude ratios of patient G’s saccades were similar to the control data.

**Figure 8.**
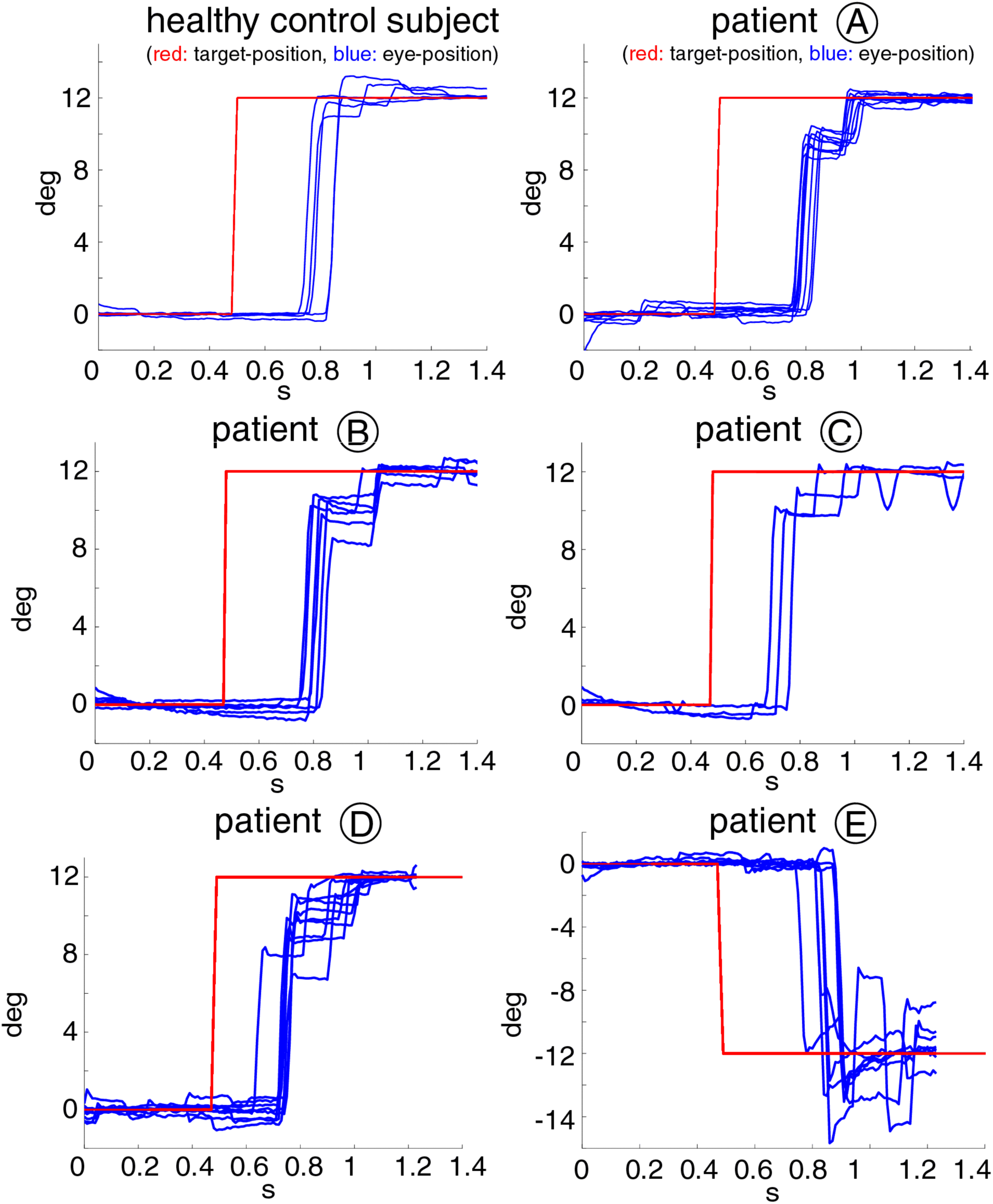
Saccadic dysmetria of the PN patients compared to saccdes of a healthy control person. The saccades performed by the PN patients were predominantly hypometric (patients A to D), in one case (patient E) hypermetric. Left upper panel: saccades

**Figure 9.**
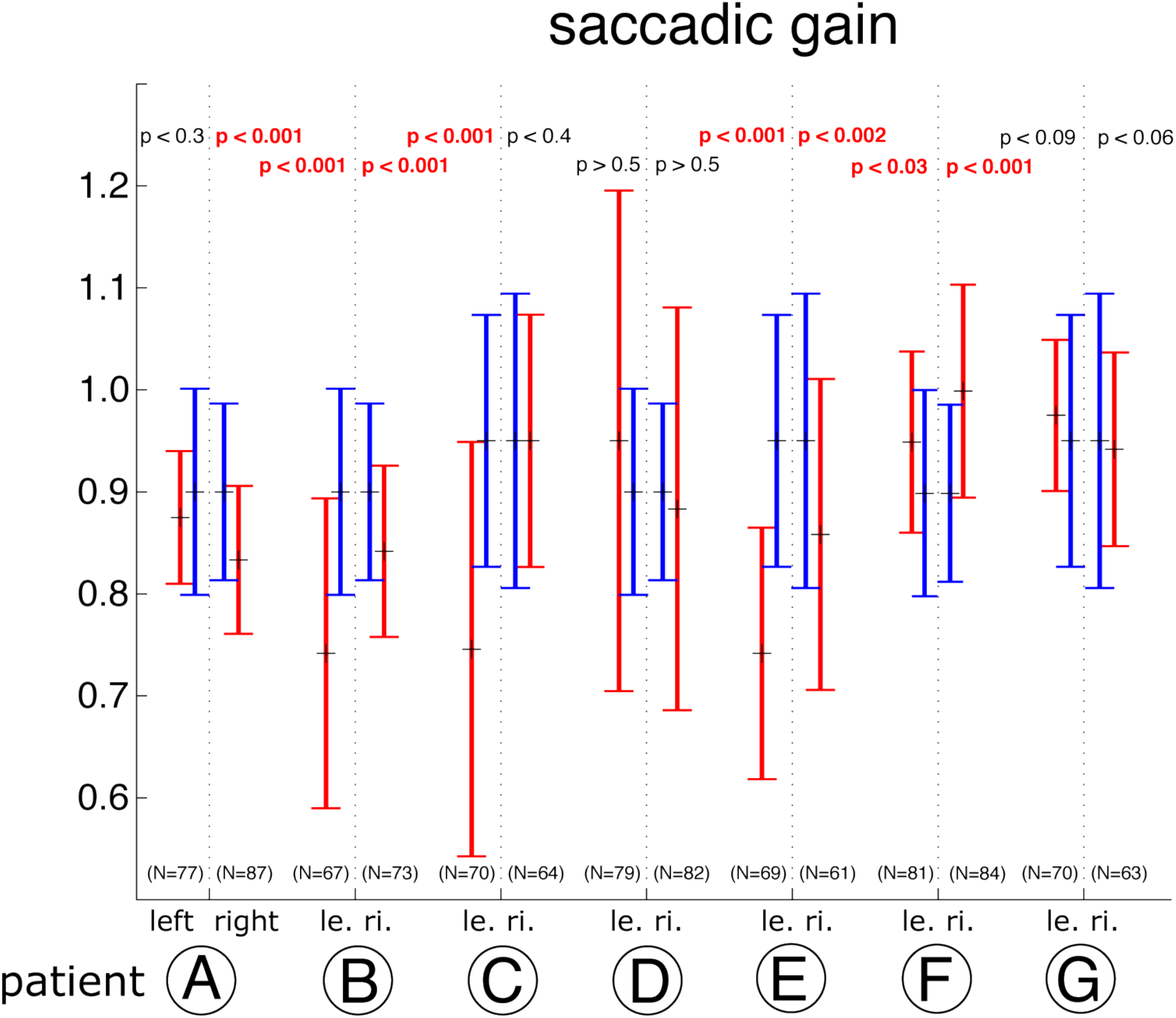
Reduced saccadic gain. The gain of left- and rightward directed saccades of the patients (red), compared to those of the control subjects (blue). All, except patients E, F and G show significantly reduced saccadic gain in at least one gaze direction.

**Figure 10.**
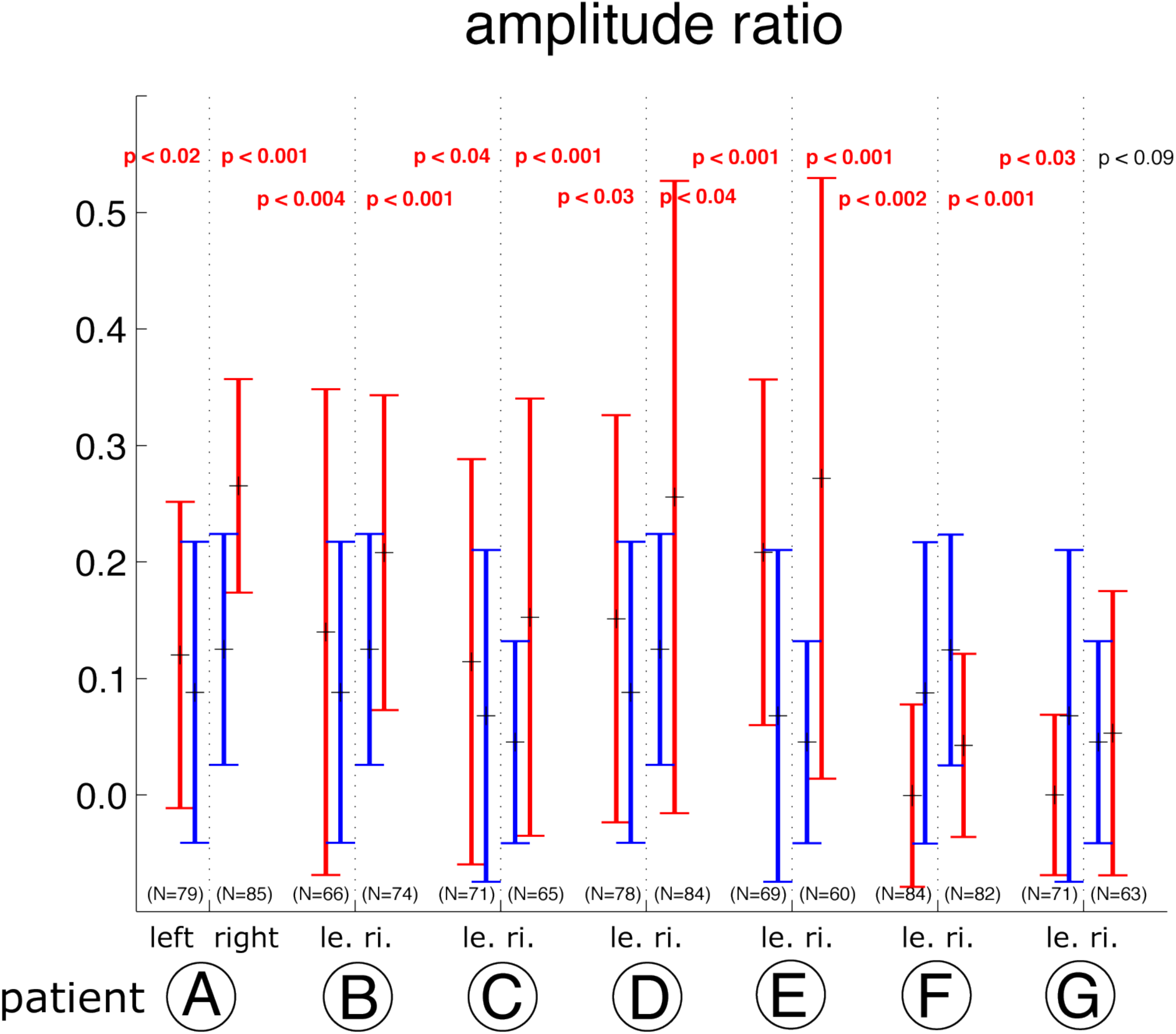
Larger corrective saccades. The corrective saccades of PN patients A to E (red) had a mean amplitude of approx. 20% of the initial saccades’ size. Patient F showed smaller corrective saccades than the controls. For the controls and patient G, the amplitude ratio was at approx. 10%.

No significant differences could be found between the latencies or durations of control and patient saccades.

### Saccades: main sequence

Disturbances of the dependency of maximum eye velocity or duration from saccade amplitude as represented by the saccadic main sequence could point to deficits in low-level saccadic mechanisms. A paresis of eye muscles e.g. would decrease the maximum velocity for a given amplitude and consequently the maximum velocity to amplitude ratio would be smaller than for the contols. Figure 11 shows, that the course of the main sequences of controls (blue) and patients (red) is identical. Although the patients’ amplitudes are smaller, the maximum velocities and durations scale to fit the main sequence. Both normal and patient saccades show a nonlinear saturation effect above 8 deg which has previously been described (Leigh and Zee, 1991). The patients’ ratio between maximum eye velocity and saccade amplitude is not different from the one of the controls while the gain of the saccades is reduced (Figure 12). These findings indicate that the patients’ saccadic dysmetria is not due to a low-level deficit but rather a deficit in the higher level planning and/or coordination of the eye movement.

**Figure 11.**
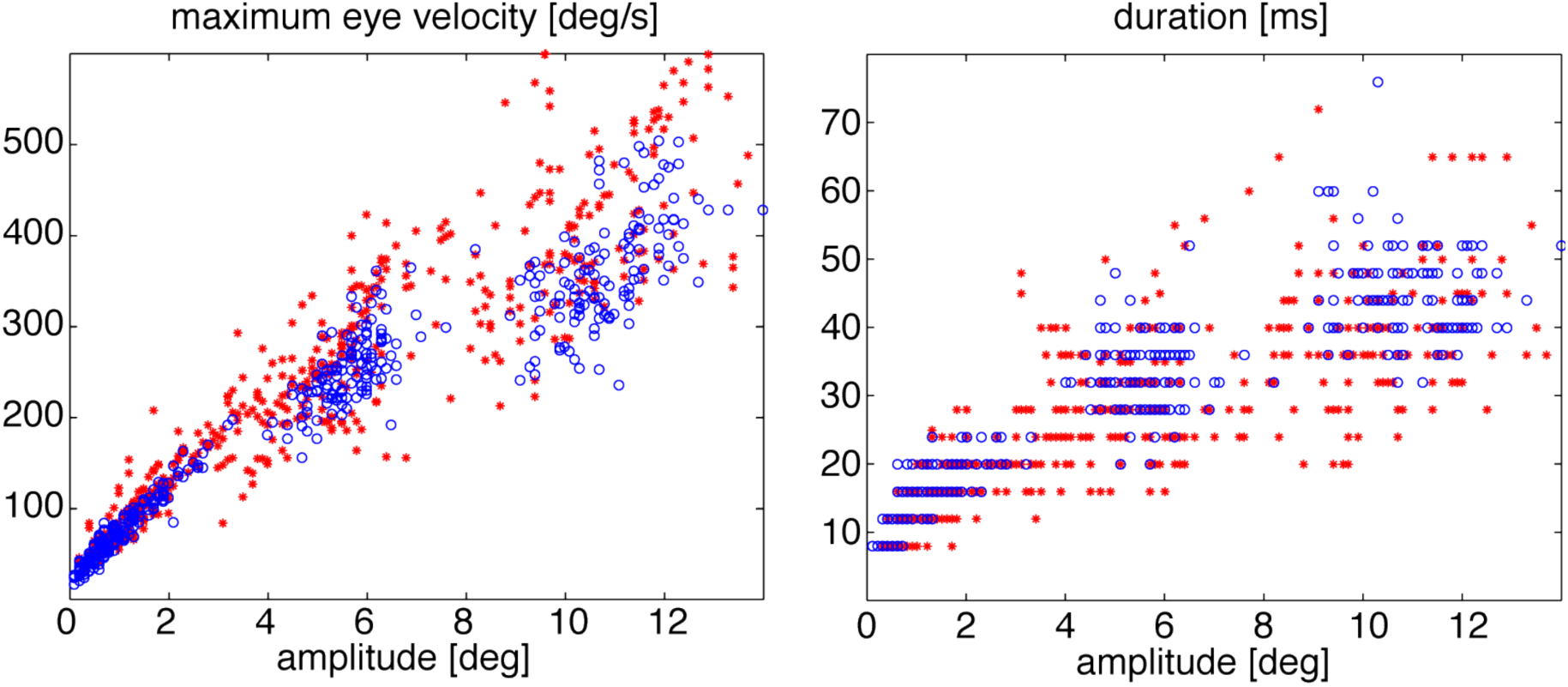
Saccade main-sequences. red: PN-patients, blue: corrensponding control subjects left: saccadic maximum velocity as a function of amplitude right: saccadic duration as a function of amplitude The clusters correspond to corrective saccades and saccades to targets at ±6 and ±12 deg respectively. Although the amplitudes of the patients’ saccades are reduced, the course of the main sequences is unaffected.

**Figure 12.**
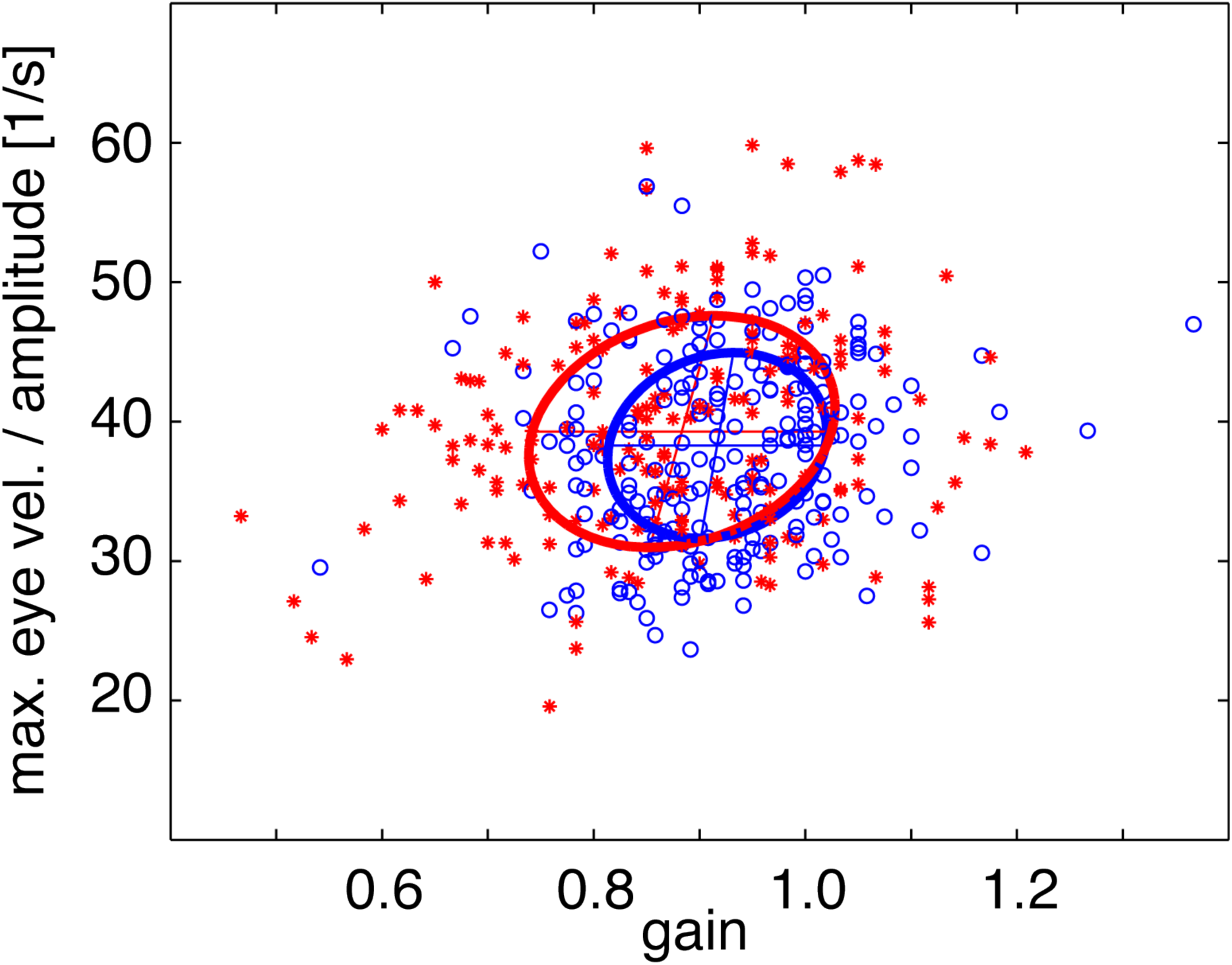
Normalised maximum velocity. red: PN-patients, blue: corrensponding control subjects The quotent of maximum velocity and amplitude is the normalised maximum velocity. Despite of smaller gain for the patients it is statistically identical for the two groups. For this analysis only the initial saccades were chosen.

### Skewness

Van Opstal and van Gisbergen (Van Opstal and van Gisbergen, 1987) have pointed out, that the ratio between the eye movement acceleration and deceleration time interval ist not constant for all saccades. Such deviations from a symmetrical, bell-shaped velocity profile occur especially in saccades with large amplitudes or as a result of fatigue (Collewijn et al., 1988). Van Opstal and van Gisbergen introduced the parameter skewness as a measure for this property of the eye velocity profile. The skewness of saccade velocity profiles can be used to reveal the underlying mechanisms of saccade preparation and control. The control subjects’ saccadic velocity profiles show a correlation between skewness and duration (Figure 13, left, blue regreesion line, Table 1). Longer saccades are more skewed: their deceleration time exceeds the acceleration time about up to 50%. Short saccades are not completely symmetric as well. Their mean skewness has a value of approximately 10%.

**Figure 13.**
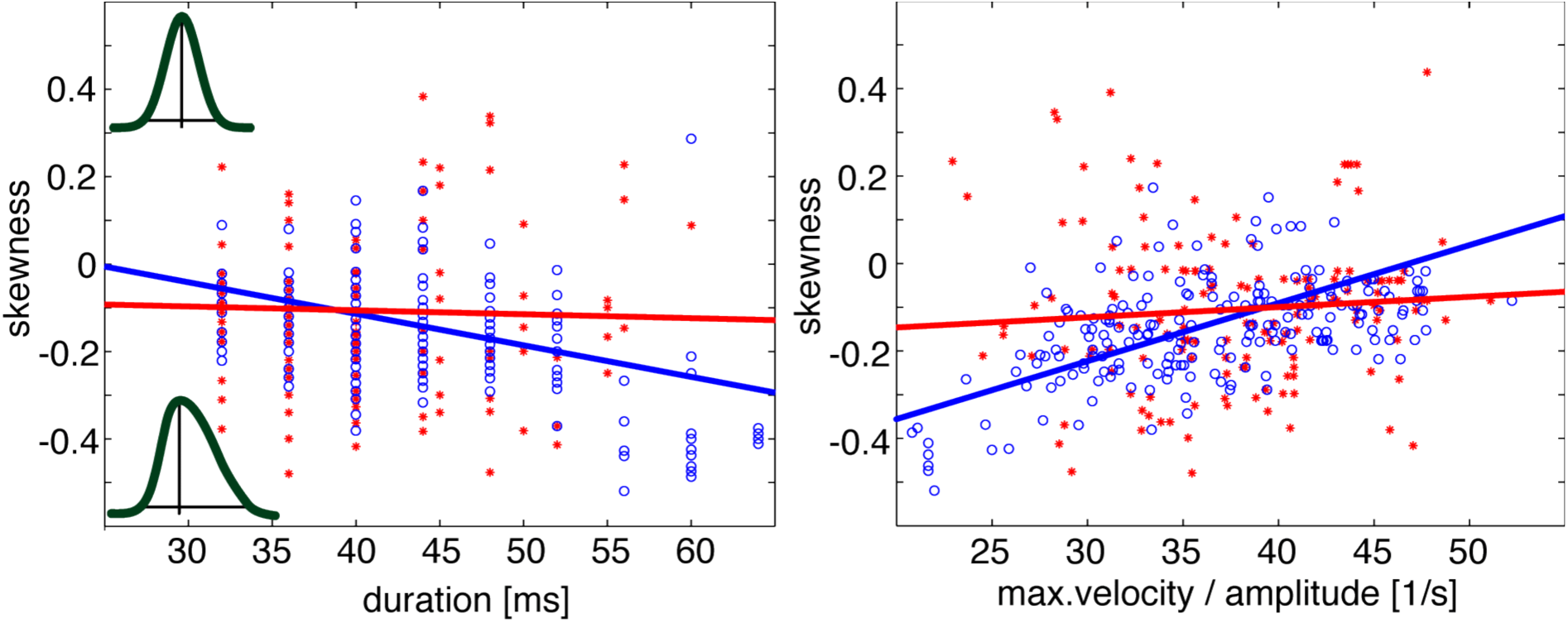
Dependency of measured skewness from saccade duration and normalised maximum velocity. red: PN-patients, blue: corrensponding control subjects left: For longer saccades the skewness of the healthy control subjects becomes negative and larger (blue regression line) which represents a less symmetric velocity profile (inlays). right: Saccades of healthy control subjects, that have a higher maximum velocity at a given amplitude (a higher normalised maximum velocity) are less skewed than the relatively slower saccades (blue line). The hypometria caused by the smaller velocity is therefore compensated by a prolongation of the deceleration phase. Both relationships no longer persist at the PN patients (red lines, see Table 1).

**Table 1:** results of linear regression (saccades) The coefficients a_0_ and a_1_ result from a linear least squares fit to y = a_0_+ a_1_x. r: correlation coefficient, p: p-value of the correlation pat.: PN patients, ctr.: control subjects, v_max_: maximum saccade velocity, v_max,norm_: normalised maximum saccade velocity The correlations of skewness and saccade duration and normalised maximum saccade velocity broke down for the patients (gray fields).

| correlation between | a<sub>0</sub><br>(pat.) | a<sub>1</sub><br>(pat.) | r<br>(pat.) | p<br>(pat.) | a<sub>0</sub><br>(ctr.) | a<sub>1</sub><br>(ctr.) | r<br>(ctr.) | p<br>(ctr.) |
| --- | --- | --- | --- | --- | --- | --- | --- | --- |
| v<sub>max</sub> - amplitude | 61,4 | 34,3 | 0,84 | <0,0001 | 45,1 | 27,9 | 0,81 | <0,0001 |
| v<sub>max</sub> - amplitude<br>(catch-up saccades) | 17,2 | 37,9 | 0,94 | <0,0001 | 14,4 | 41,0 | 0,90 | <0,0001 |
| duration - amplitude | 13,4 | 2,6 | 0,60 | <0,0001 | 12,7 | 3,02 | 0,85 | <0,0001 |
| skewness - duration | -0,07 | -0,001 | **0,002** | **59,1** | 0,17 | -0,007 | 0,15 | <0,0001 |
| skewness - v<sub>max,norm</sub> | -0,20 | 0,002 | **0,009** | **24,1** | -0,62 | 0,013 | 0,54 | <0,0001 |

The healthy subjects additionally exhibit a correlation between maximum velocity normalized on amplitude and skewness (Fig. 13 right, blue line, Table 1). The skewness is negative and smaller for slower saccades indicating a more symmetrical velocity profile and a shorter deceleration time for the faster saccades at a given amplitude.

Figure 14 makes clear that normal saccades with higher absolute skewness (red, S < −0.1) for any measured amplitude have a decreased maximum velocity relative to the symmetrical ones (blue, S > −0.1). The smaller saccades (to targets at ±6 deg) are more symmetrical in their velocity profile than the larger ones (to targets at ±12 deg), as indicated by the higher portion of blue points. Nevertheless, fitting a two-dimensional third order polynomial function to the data results in a skewness surface clearly descending towards smaller maximum velocities for all amplitudes.

**Figure 14.**
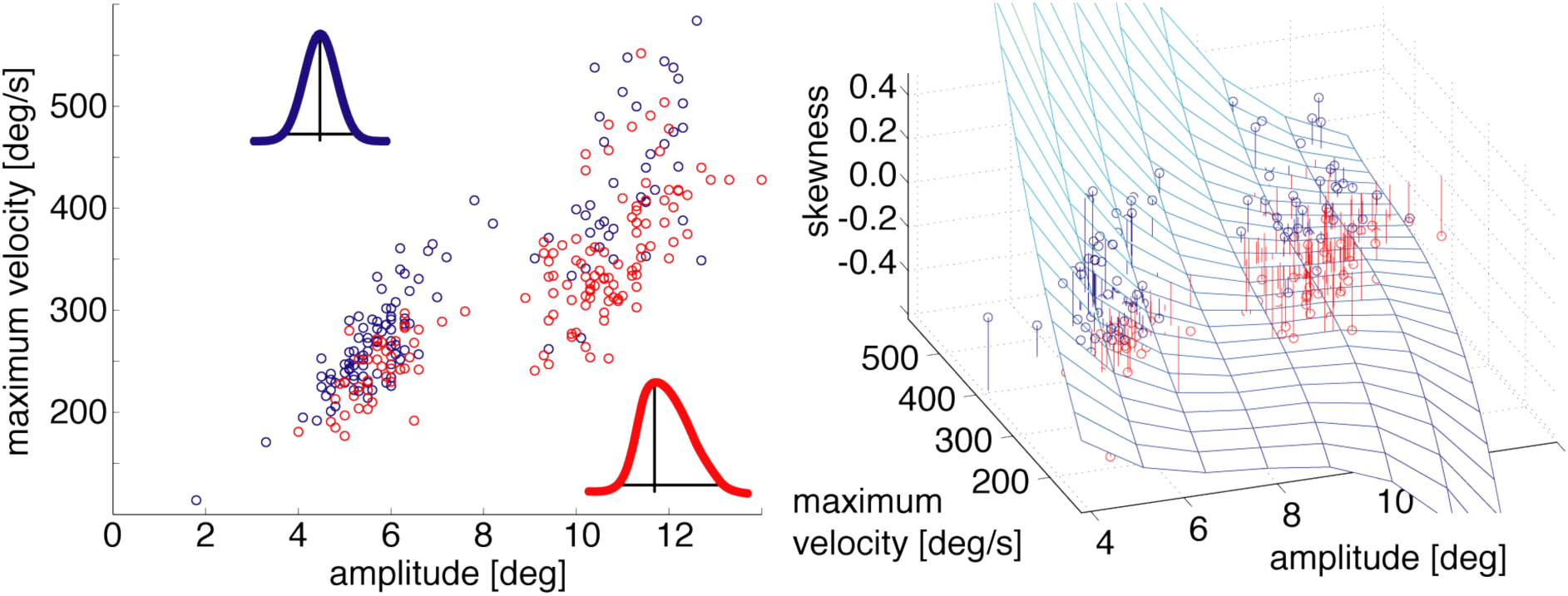
More skewed saccades have a smaller maximum velocity. Saccades with a larger negative skewness in their velocity profile (red, S < −0,1), have a smaller maximum velocity than the more symmetric saccades (blue, S > −0,1). Larger saccades are more skewed (more red dots), but the relationship with the maximum velocity persists along the whole main sequence (right, fit of a plane, defined by a 3^rd^ order two-dimensional polynomial)

Both correlations, skewness vs. duration and skewness vs. normalized velocity do no longer persist in the saccades performed by the PN patients (Figure 13, red line, Table 1). Their skewness varies for all measured saccade duration values over the entire range of −40% to 40%.

Table 1 summarizes the results of linear correlation calculations for the saccadic main sequences and the dependencies of skewness on saccade duration and on maximum velocity per amplitude. While with the main sequences a good consistence between patients and controls prevails, the skewness dependencies on duration and maximum velocity per amplitude are no longer significant within the patient data.

In order to determine, whether for the skewness an interaction between the factors group affiliation (patients resp. controls) and duration and/or maximum velocity per amplitude exists, a two-factorial ANOVA was computed. The results (table 2) indicate the absence of such an interaction.

**Table 2:** results of a two-factorial ANOVA. For the dependency of saccade skewness from duration and from normalised maximum velocity as well there exists no interaction between group membership (patiens or controls) on one hand and duration/normalised maximum velocity on the other (gray fileds).

| Factor 1 | Factor 2 | p-value<br>contribution of factor 1 | p-value<br>contribution of factor 2 | p-value<br>interaction |
| --- | --- | --- | --- | --- |
| group (pat./ctr.) | duration | 0,16 | < 0,05 | 0,56 |
| group (pat./ctr.) | $V_{\max, \text{norm}}$ | 0,73 | < 0,05 | 0,90 |

### Smooth pursuit eye movements

As mentioned before, the participation of the PN in the execution of SP eye movements has previously been shown by several studies using e.g. electrophysiological methods on monkeys and cats as well as studies on five human patients with pontine lesions. The examined patients of our study also showed a clear SP deficit. Examples of SP eye velocities after saccade elimination and filtering are shown in Figure 15. PN patient B (right panel) exhibited deficits in SP indicated by a lower SP gain compared with that of a healthy control person (left panel).

**Figure 15.**
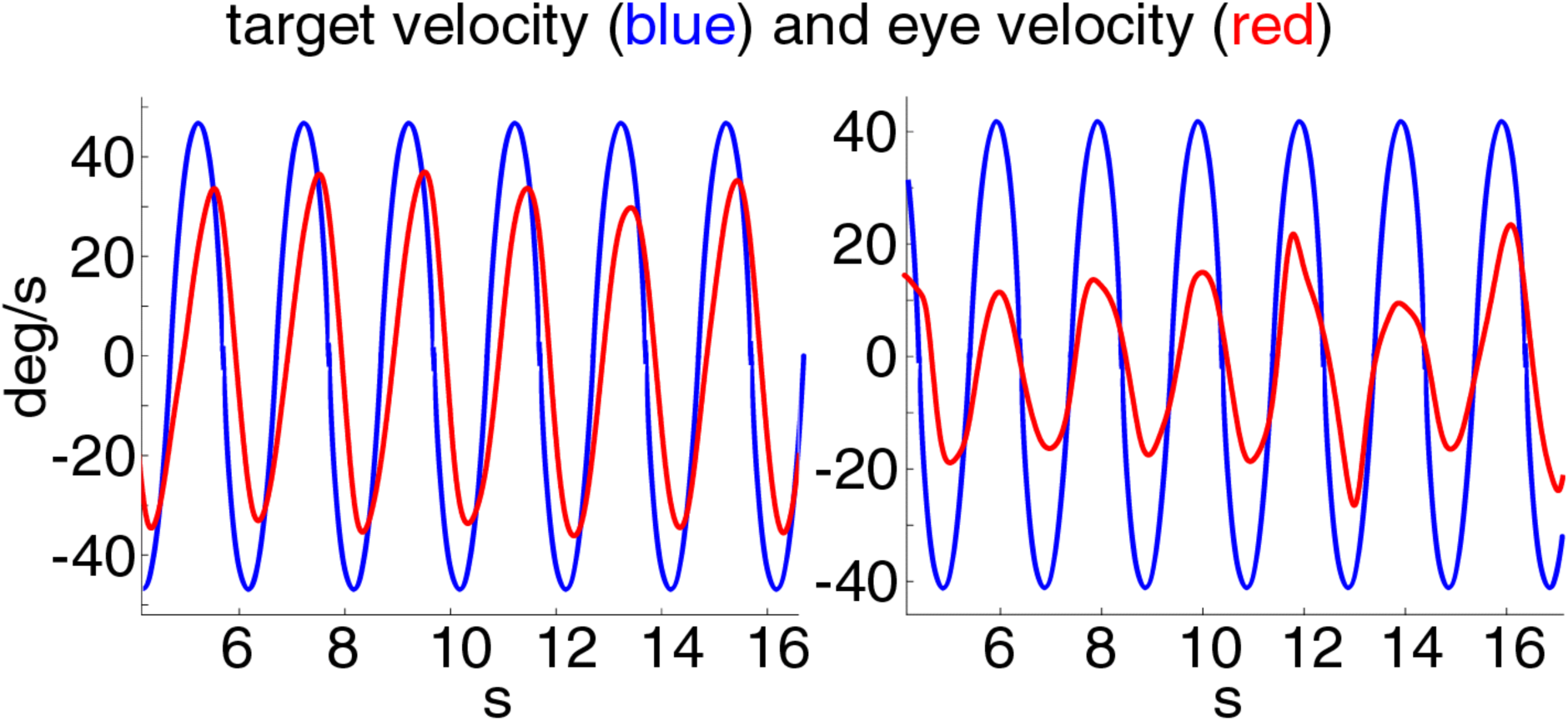
Eye-velocity after saccade elimination. Red: eye velocity, blue: target velocity left: healthy control subject, right: PN patient B. The maximum and minimum eye velocities of the patient are obviously smaller than those of the control subject.

SP eye movements were recorded with PN patients A, B, D, E and F. Therefore the SP data for these patients are compared with those of the appropriate control groups (Figure 16). A lateralization could be determined only for patient A. Ipsiversive eye movements in the direction of the lesion were normal, contraversive movements however showed a reduced gain. PN patients B, D and F exhibited a reduced gain for both directions. A tendencial but not significant decrease in gain was found at PN patient E. The patient with the medulla oblongata lesion G showed normal smooth pursuit with a mean gain of 1.

**Figure 16.**
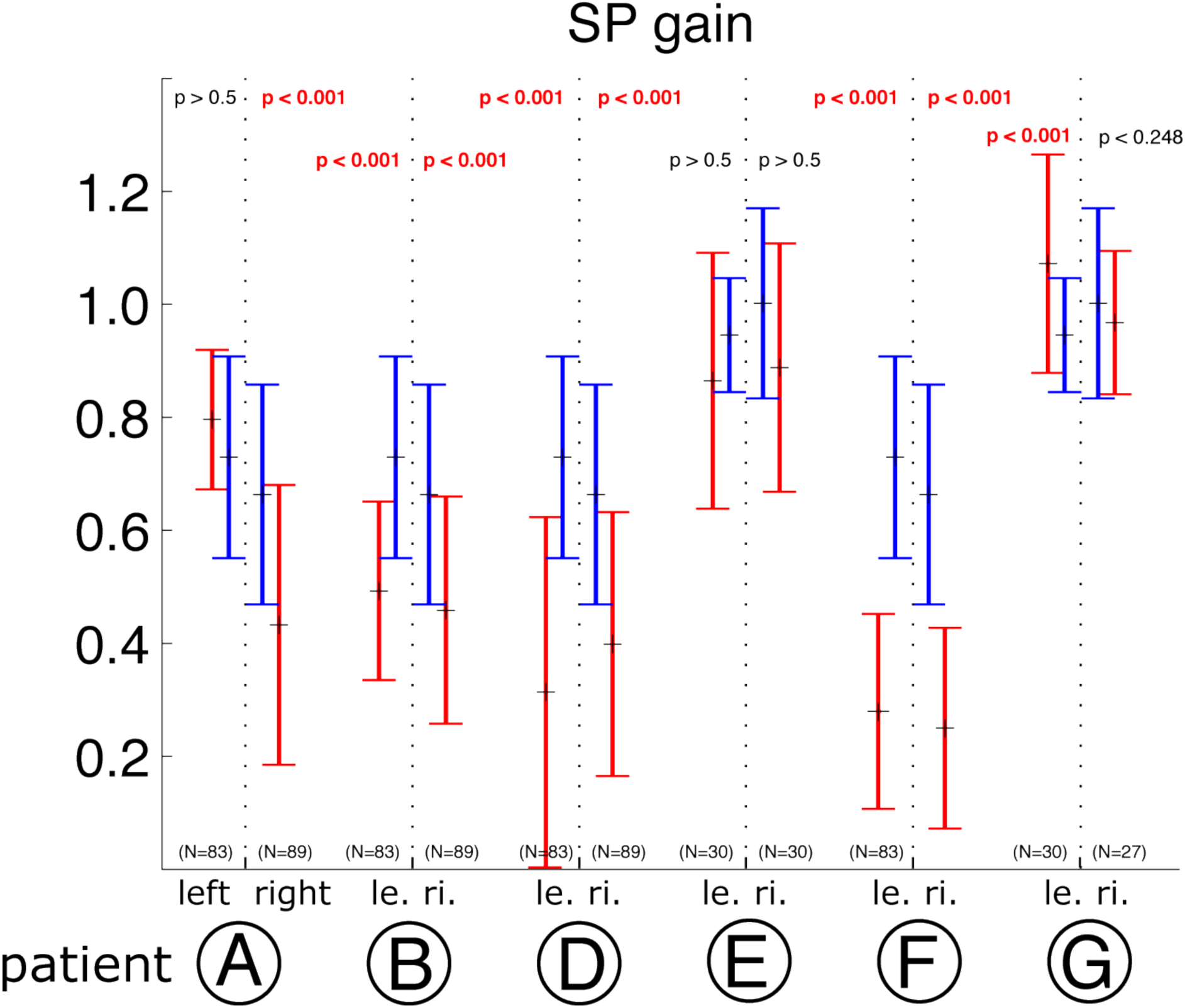
Smooth pursuit gain of patients A, B, D, E, F and G. red: patients, blue: corrensponding control subjects The SP gain of the PN patients A, B, D and F is reduced compared with the control subjects’ (A contralaterally). PN patient E and medulla patient G showed SP with normal gain.

### Pointing movements

The pointing movement trajectories of all PN patients except patient F featured an increased spatial curvature compared to those of the healthy controls (see examples in Figure 17). Therefore the maximum perpendicular distances from index fingertip to the line connecting movement start and end point (beeline) were significantly increased (Figure 18). PN patient A showed a slightly but significantly increased curvature with the movements of the contralateral arm (lesion left-sided). A clearly bilateral increase with a more ipsilateral accentuation was obvious at PN patients B, C, D and E. Movements with the right hand were normal in Patient F. All measured mean pointing movements of the PN patients except patient F were therefore higher curved than those of the corresponding controls. The pointing movements of patient G were normal in curvature.

**Figure 17.**
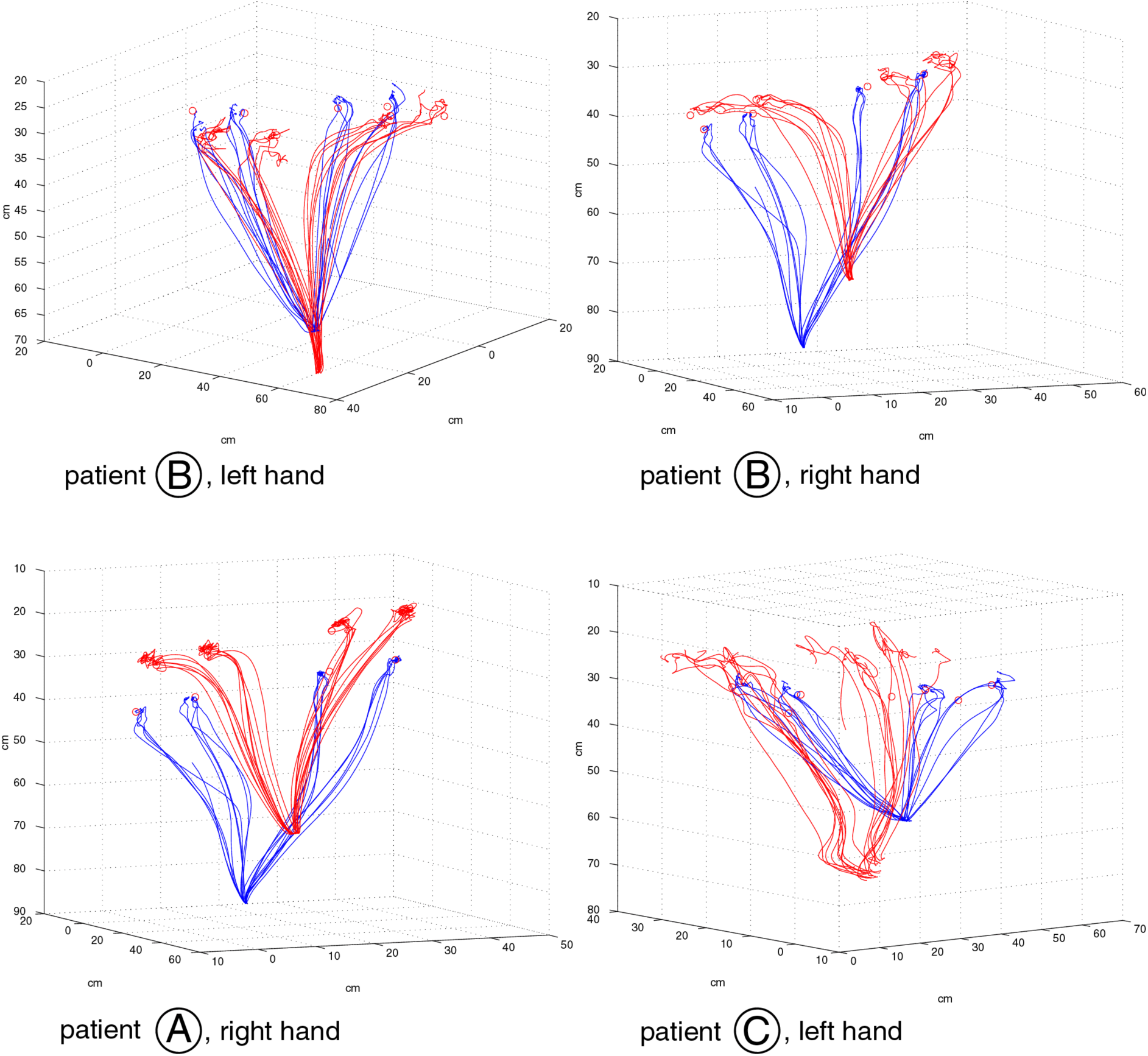
Forefinger positions during pointing movements of patients (red) and a healthy control subject (blue) towards 4 visual targets. Upper panels: PN patient B, left: pointing with left hand, right: pointing with right hand Lower left panel: PN patient A, right hand; lower right panel: PN patient C, left hand

**Figure 18.**
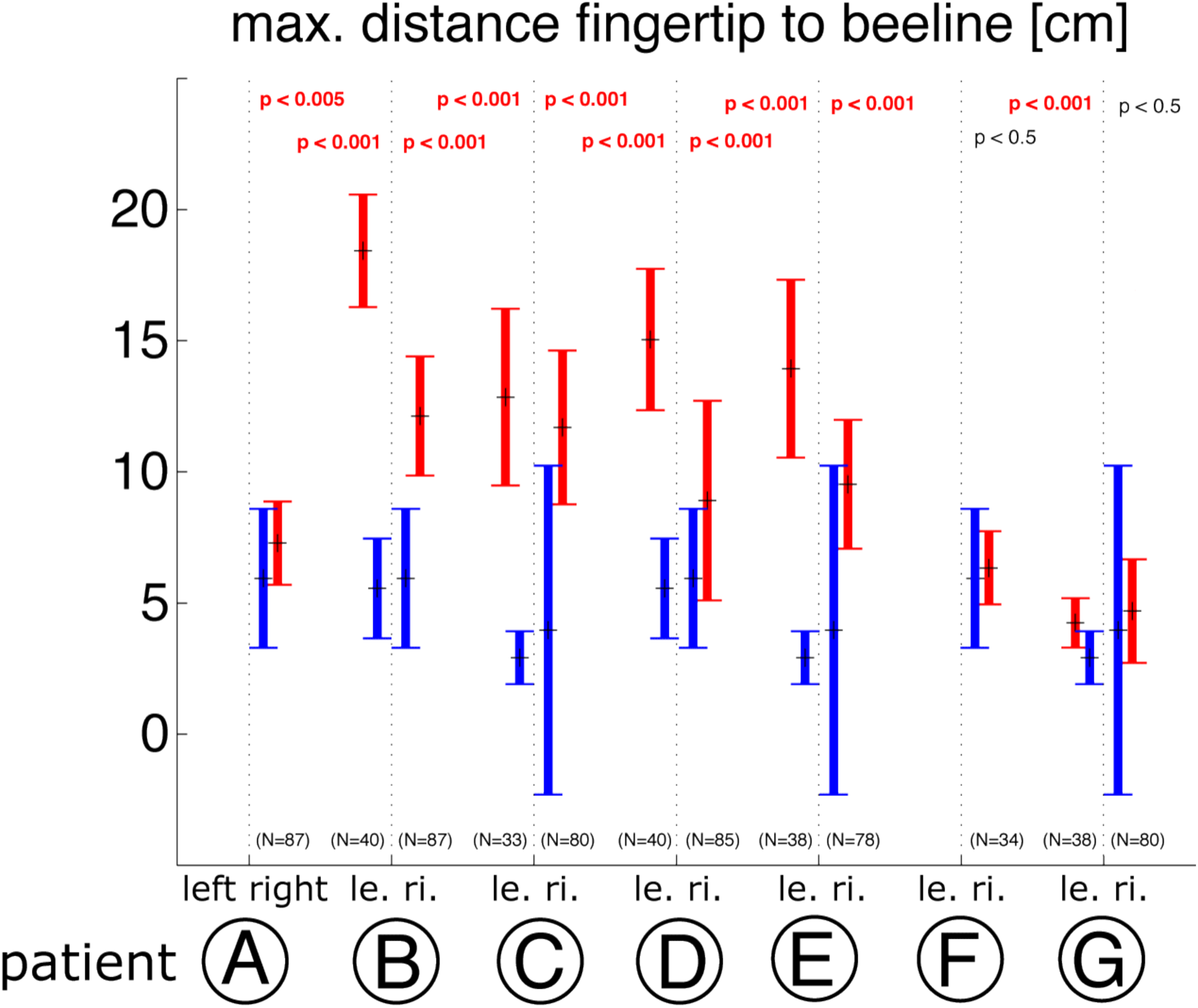
Maximum perpendicular distance to beeline as a measure for pointing movement curvature. All PN patients except patient F showed more curved pointing movement trajectories compared to the movements of the corresponding control subjects. Patients F and G performed normally curved poiting movements.

Only patients C (bilateral) and D (right arm) exhibited a significantly increased position error at the end of the movement. All other patients performed the pointing with normal accuracy.

The deceleration phase of the normal pointing movements was 2 to 3 times longer than the acceleration phase (skewness 1/3 to 1/2). Patients B and D showed bilaterally a prolongated deceleration to 3 to 4 times the acceleration time (skewness 1/2 to 3/5). The right hand pointing movement velocities of patient C were significantly more skewed as well.

Main sequences of the pointing movements were calculated. The correlations between movement duration and amplitude and between maximum velocity and amplitude were significant for both PN-patient and control data, although the patients’ amplitudes were clearly enlarged (Figure 19, Table 3).

**Figure 19.**
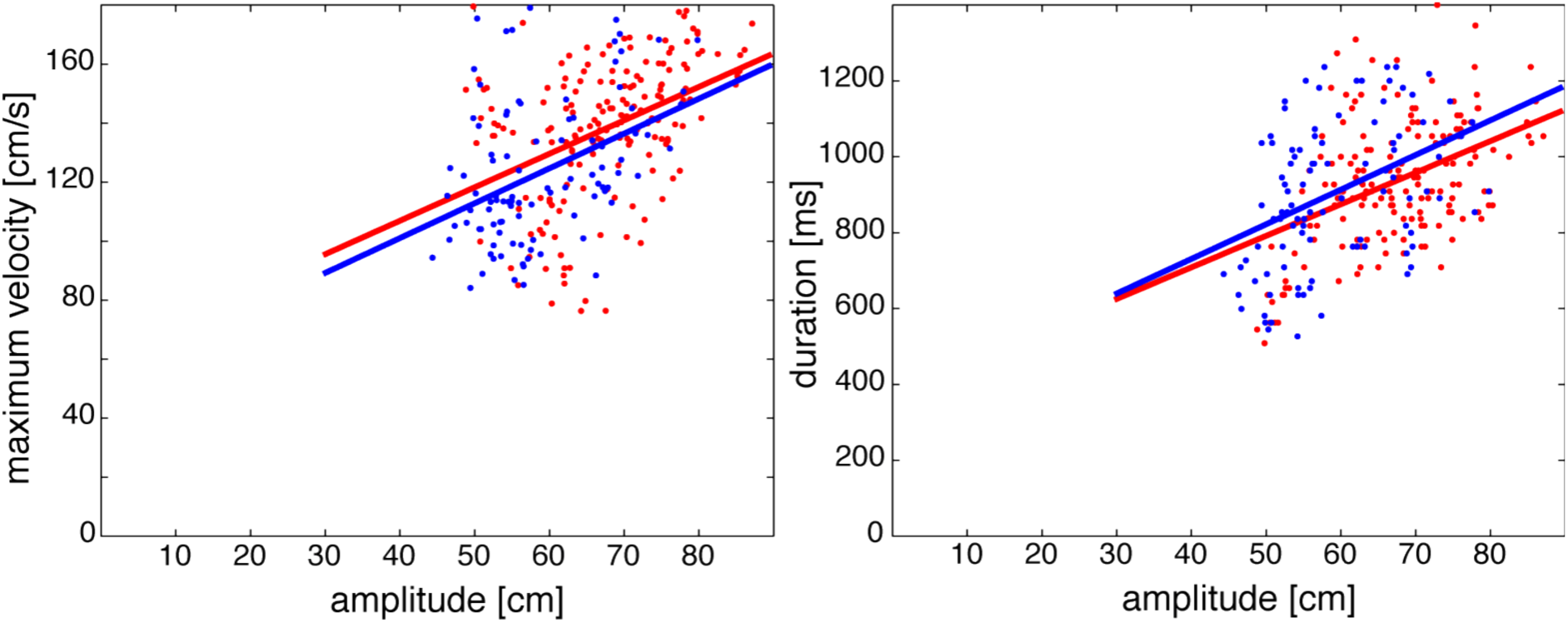
Pointing movement main sequences. red: PN-patients, blue: corrensponding control subjects right: maximum velocity of the pointing movement as a function of its amplitude left: pointing movement duration as a function of its amplitude The main sequence course is comparable for patients and controls although the amplitudes of the patients’ movements are enlarged.

**Table 3:**
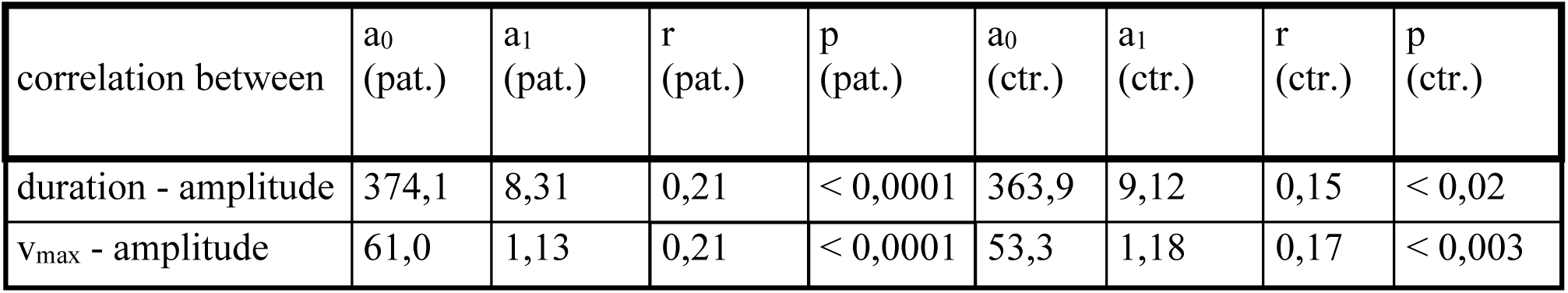
results of linear regression (pointing movements) The coefficients a_0_ and a_1_ result from a linear least squares fit to y = a_0_ + a_1_ x. r: correlation coefficient, p: p-value of the correlation pat. = PN patients, ctr. = control subjects, v_max_ = maximum velocity

## Discussion

On examining and quantifying the consequences of lesions of the human pontine nuclei (PN), we found deficits in saccadic eye movements, smooth pursuit eye movements (SP) as well as pointing movements. 5 of the 6 PN patients showed disturbances in saccade metrics (2 contralaterally), 4 of 5 PN patients exhibited SP deficits (one of these contralaterally) and 4 of 6 PN patients performed pointing movements with bilaterally increased curvature. These findings support the hypothesis, that the PN as a part of the cortico-ponto-cerebellar pathway play an important role in the generation and coordination of different types of movements.

The functional and anatomic results are summarized in Figure 20. Nissl-stained histological sections of the human PN at levels 1 to 5 (shown in the lower right panel in an exemplary saggital MRI section, from caudal to rostral) are depicted. Colored lines indicate borders of the lesioned areas, each patient in different color. At the bottom of the figure, colors underlying the labels correlate the patients to the lesioned areas depicted in the left column. The numbers indicate the sections in which tissue was lesioned. Figure 21 shows a table, in which for every patient (rows) and every type of movement (columns: saccades, SP, pointing) the experimental findings and (if available) their lateralization is summarized. Results are grouped in the categories deficits (saccadic hypometria or increased variability [patient D], reduced SP gain or increased curvature of pointing trajectory, respectively), normal movements and not measured movements. Lateralization is indicated by letters (i=ipsilateral, c=contralateral, b=both sides) which are set in brackets in case of an unilaterally higher effect.

**Figure 20.**
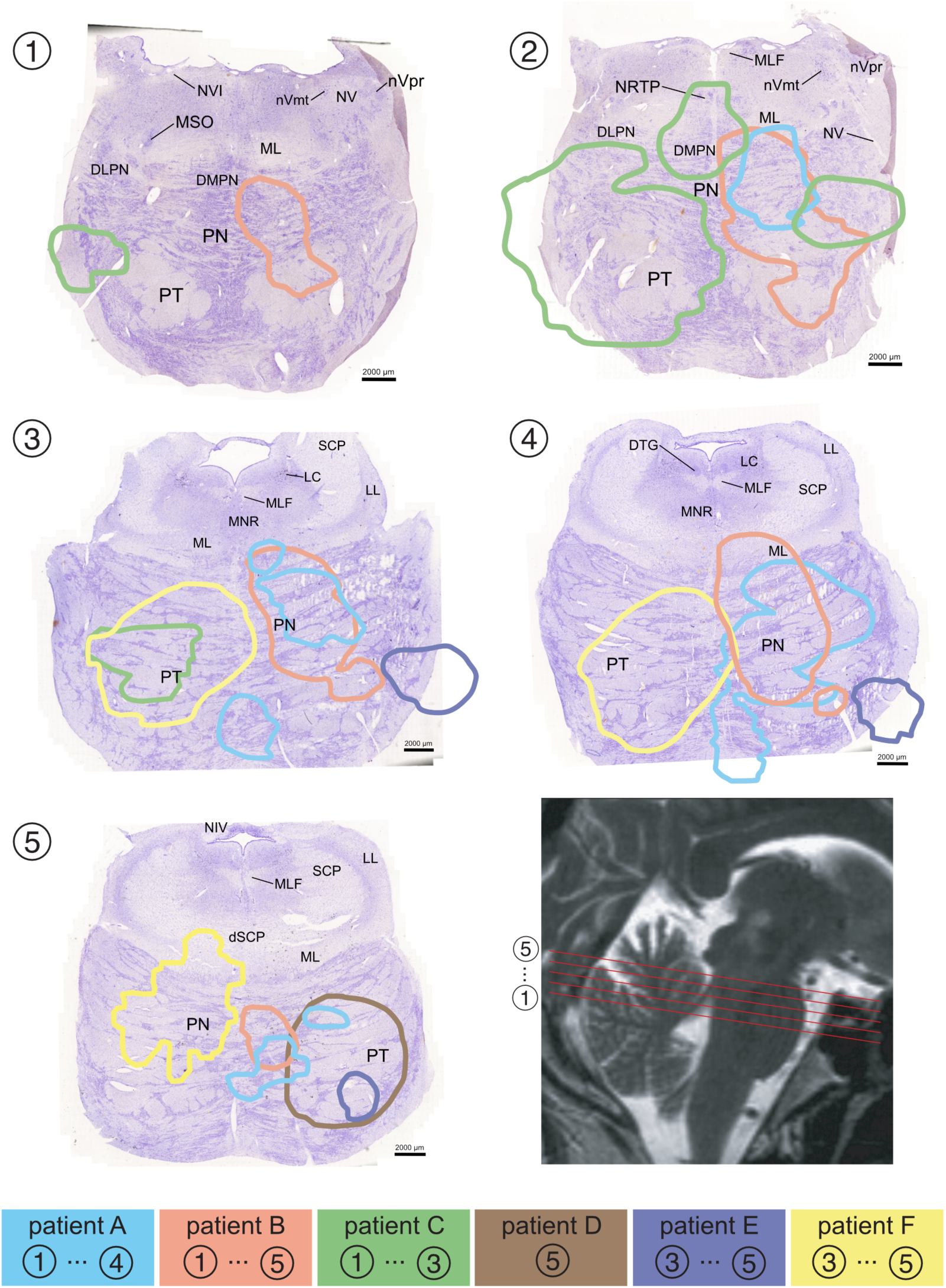
Anatomical location of PN patients’ lesions. Panels 1 to 5: Histological sections of the human pontine Nuclei (Nissl staining) at levels 1 to 5 (from caudal to rostral). The colored lines illustrate the lesioned areas of the patients, each in different color. Lower right panel: The location of the corresponding section levels drawn into an exemplary sagittal MRI image. Bottom line: The colors correlate the patients to the lesioned areas shown in panels 1 to 5. The numbers indicate the section ranges in which tissue was lesioned. Labels in histological sections: DLPN: dorsolateral pontine nuclei DMPN: dorsomedial pontine nuclei DTG: dorsal tegmental nucleus LL: lateral lemniscus ML: medial lemniscus MLF: medial longitudinal fascicle MNR: median raphe nucleus MSO: medial superior olive nVmt: motor trigeminal nucleus nVpr: principal (sensory) trigemnal nucleus NV: trigeminal nerve NRTP: nucleus reticularis tegmenti pontis PN: pontine nuclei PT: pyramidal tract dSCP: decussation of the SCP SCP: superior cerebellar peduncle

**Figure 21.**
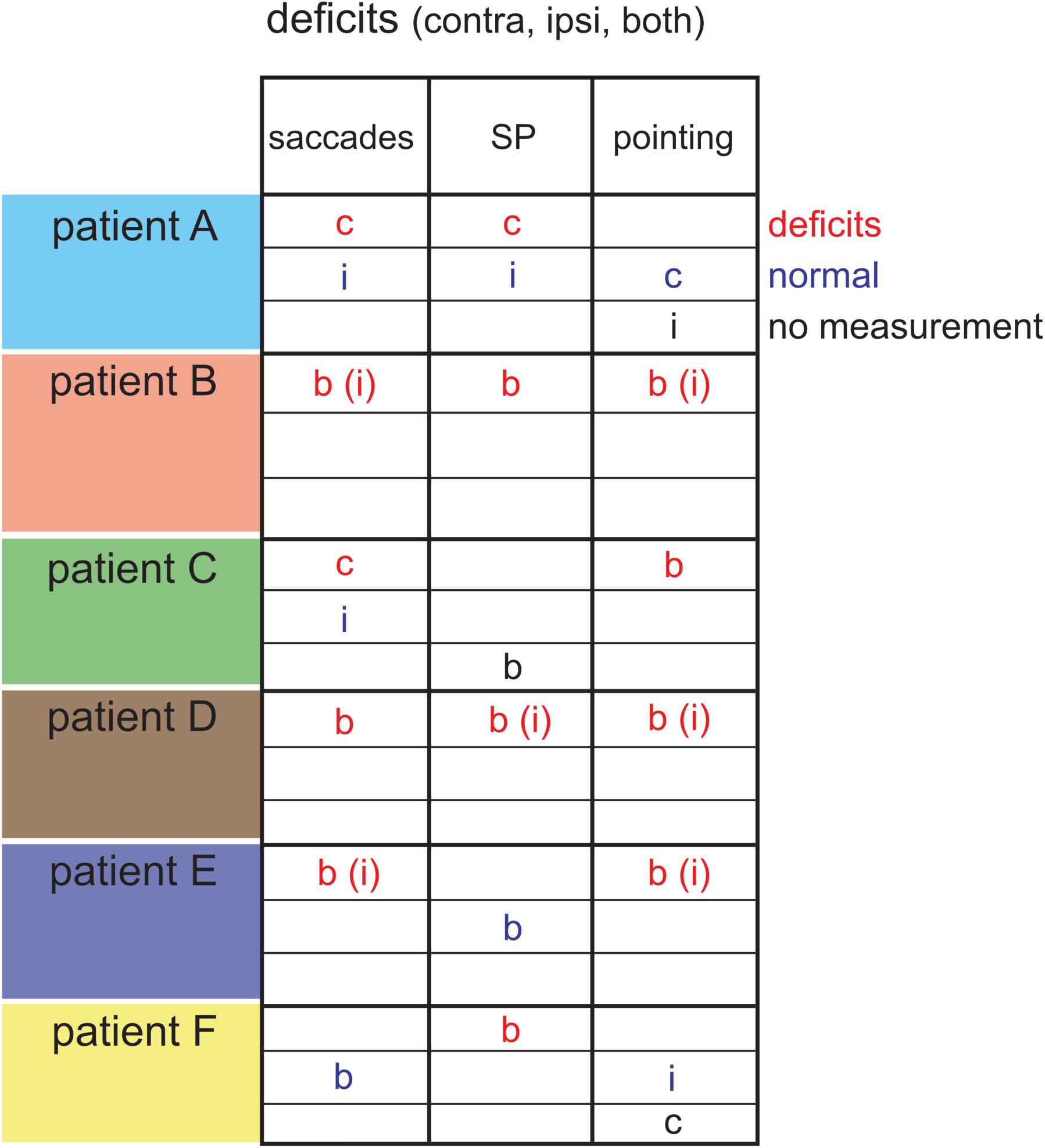
Summary of PN patients’ movement deficits. For every patient (rows) and every type of movement (columns: saccades, SP, pointing) this table shows the experimental findings and (if available) their lateralization. Results are grouped in the categories deficits (saccadic hypometria or increased variability [patient D], reduced SP gain or increased curvature of pointing trajectory, respectively), normal movements and not measured movements respectively. Lateralization is indicated by letters (i=ipsilateral, c=contralateral, b=both sides) which are set in brackets in case of a higher effect unilaterally.

Our results clearly demonstrate, that lesions of the human pons, besides the well known SP deficits, do additionally impair visually guided saccades. We could show that the pontine lesions of patients A, B, C and E caused hypometric saccades for at least one gaze direction and an increased saccade amplitude variabililty for patient D. Patient F showed a saccadic hypermetria of ipsilateral corrective saccades. Patient A exhibited a reduced saccadic gain and bigger corrective saccades for eye movements to the right (contraversive). A lateralization was obvious in the saccadic deficits of patient C (leftward, contraversive movements) as well, while the deficits of patients B, D and E were bilateral.

It has previously been shown (Duhamel et al., 1992; Li et al., 1999), that contraversive saccades are disturbed in the presence of cortical lesions. If the patients’ pontine lesions had affected the cortico-ponto-cerebellar pathway, then contraversive saccade deficits should have been caused. In fact, the two patients A and C whose saccadic deficits showed a lateralization both suffered from contraversive saccadic deficits. It is possible, that here more PN cells involved in oculomotorics than crossing ponto-cerebellar fibers were affected. This finding could be due to the spreading of the ponto-cerebellar fibers after crossing the side as described by Schmahmann and co-workers (Schmahmann et al., 2004b).

The patient presented by Thier et al. (Thier et al., 1991) had a lesion that aligned with patient D’s lesion. They reported a hypometria in 50% of ipsiversive and 22% of contraversive saccades. Our patient D however, only showed an increased gain variability. The reason for this difference might be the fact, that patient D’s lesion was caused by a tumor that kept intact some of the neurons in the affected area. On the other hand, we could reproduce their finding of a bilateral SP deficit. Ipsi- and bilateral SP deficits caused by PN lesions were as well reported by Gaymard et al. (Gaymard et al., 1993). Our results for the first time quantitatively provide evidence for saccadic dysmetria (especially hypometria) caused by lesions in the human PN.

In direct neighborhood to the PN lies the tegmentum from which the rectus lateralis innervation via the abducens nerve emerges. A lesion affecting the tegmentum would cause eye movement deficits as well. Patient 2 in Gaymard’s study (Gaymard et al., 1993) showed a lesion clearly extending into the tegmentum which yielded saccades that were hypometric and had a lower maximum velocity. Eye muscle paresis leads to a reduced quotient of maximum velocity and amplitude for directions affected by the paretic muscle (Metz, 1983). A tegmental lesion would therefore have caused saccades with clear differences in the main sequence. Our finding that the PN patients’ main sequences were unaffected (Figures 11 and 12, Table 1) strongly suggests that the lesions of our PN patients did not extend into the PPRF or NRTP. The higher SP gain of patients E and G and of the correlated controls compared to those of the other patients (e.g. A, B or D) can be explained by the lower age of patients E and G. The age-dependency of SP gain has been shown previously (Morrow and Sharpe, 1993).

Patients E and F showed a functional segregation of the lesions’ effects on different types of eye movements, because saccades were affected and SP remained intact and vice versa. The more ventro-laterally located smaller lesion of patient E could have affected different oculomotor patches of PN neurons than patient F’s lesion. Whether the distribution of oculomotor functions in the PN is really inhomogeneous is yet unclear. On one hand it is supported by the anatomical studies of Schmahmann et al. (Schmahmann et al., 2004c), on the other hand, electrophysiology in nonhuman primates suggests a more heterogeneous representation of eye movements (Dicke 2004, Tziridis 2009). Saccades and SP of control patient G with the medulla oblongata lesion did not differ significantly from the movements performed by the control subjects. Apparently, this lesion did not affect oculomotor structures.

Tziridis et al. (Tziridis 2009) found neurons responding to hand movements in the dorsal pontine nuclei of nonhuman primates. These neurons were generally located more medially than those responding to eye movements. In contrast our findings in patient A, with a more medial lesion and normal contraversive pointing movements and in patient E with a lateral lesion and impaired saccades and limb movements combined with intact SP eye movements challenge the view of a clear functional mapping in the PN. Over all it seems hard to easily correlate this study’s human lesions with the results from studies using electrophysiological single-unit activity measurements. Nevertheless Tziridis et al. found a large heterogeneity of the eye- and hand-movement related neuron types. They in fact point out that their results speak against an encoding of movement kinematics or dynamics or the processing of proprioceptive signals but are more in line with a representation of higher level premotor information processing.

Analyzing the velocity profile of the patients’ hypometric saccades allowed us to obtain more information on how the cerebellum might perform its task as a motor control and calibration unit. The PN patients’ lesions not only caused saccadic dysmetria, but also destroyed the dependencies of velocity skewness from saccade duration and from maximum velocity. The reason could be an impairment of calibration mechanisms supported by the cotico-ponto-cerebellar pathway.

A number of studies on nonhuman primates as well as cerebellar patients have shed some light on how the cerebellum is involved in motor learning, especially the oculomotor vermis (OMV) in short term saccade adaptation (STSA) ond oculomotor fatigue. They have been reviewed recently (Prsa and Thier 2011). In a nutshell it has been proposed that the cerebellar cortex is calibrating the timecourse of a movement by changing the amount of contributing purkinje-cells via adjustment of the parallel-fiber-purkinje-cell-synapses. The purkinje-cell single-spike activity shows an extended deceleration phase for larger saccades. The error signal for motor learning has been assumed to be provided by the inferior olive via climbing fibers and influencing via long-term depression (LTD) the parallel-fiber-purkinje-cell-synapses.

Recent work (Junker et al. 2018) has callenged this view and suggests a more complicated influence of the olivary signal. Sun and co-workers (Sun 2017) showed the separation of adaptation-mechanisms of saccades on one hand and SP on the other in the OMV of nonhuman primates suggesting a separation of saccade- and SP-related synapses on P-cell dendritic trees.

The hypothesized mechanism of cerebellar contribution to motor learning has meanwhile been well supported by several studies including e.g. cerebellar patient studies from Golla et al. (OMV, Golla 2008) and Xu-Wilson et al. (cerebellar cortex, Xu-Wilson 2009) in which saccadic velocity profiles have been analysed. Following these findings, we tried to analyse the kinematics of eye- and limb-movements in this study’s patients with focal pontine lesions to get insight, if already at the level of the PN signs for the mentioned calibration mechanism by shaping the movements’ velocity profile can be found.

Our saccade data of healthy control subjects clearly demonstrate, that the saccade velocity skewness increases with increasing duration of the saccades. Furthermore, for a given amplitude, saccades with a lower maximum velocity have a longer deceleration phase than the faster ones. This dependency supports the hypothesis of a mechanism that prolongates hypometric movements by extending the deceleration phase. It allows the saccadic system to overcome deficits in the oculomotor plant caused e.g. by damage or fatigue. An increase of saccadic velocity skewness after fatigue, application of diazepam (Van Opstal and van Gisbergen, 1987) or after modification of saccadic gain by adaptation (Catz et al., 2005; Straube et al., 1997) has in fact been shown previously. A hypothetic inactivation of this calibration mechanism would cause hypometric eye movements like the saccades measured in all but one of our study’s PN patients. The PN lesions and the lesions of fibers connecting PN and cerebellum inhibit the necessary information for accurate calibration from reaching the cerebellar cortex. As a consequence it might be possible, that an adequate adjustment of the movement timing resp. the velocity profiles and their skewness to the necessary saccade duration and velocity is not achieved.

The illustrated deficits in high level movement calibration mechanisms might as well be responsible for the observed pointing movement deficits in the PN patients. The observed increase in pointing movement trajectory curvature reflects deficits in the temporal finetuning of muscle activation. Pointing movements however require a much higher complexity of activation adjustments than the relatively simple eye movements since in comparison far more muscles are involved leading to higher degrees of freedom.

Cerebellar lesions normally cause typical limb movement deficits like an intentional tremor or cerebellar ataxia. If cerebellar inputs are affected by pontine lesions, simliar limb movement deficits should occur as well. Problematic is the distinction between deficits caused by cerebellar malfunction and those due to lesions of the pyramidal tract which passes the pons in direct neighbourhood to the pontine nuclei. Our pointing movement data suggest that the limb movement deficits observed in the PN patients are rather due to timing and coordination deficits caused by cerebellar malfunction than to pyramidal tract impairment. Deficits in low-level pointing movement execution like those provoked by ataxic paresis should have caused more pronounced deviations from the normal main sequences which were not found. This suggests deficits in higher level planning and coordination deficits. Further behavioural and e.g. electrophysiological studies might be necessary to clarify the functional topology of movement processing in the PN.

Our results confirm the essential role of the human PN in the motor function of the cortico-ponto-cerebellar pathway. They support the hypothesis that the cerebellar cortex controls different types of eye and limb movements by generally adjusting the timing of muscle innervation via adaptation of the parallelfiber-purkinjecell synapses.

## Data Availability

Relevant data will be sent on e-mail request to the author.

## Acknowledgements

This publication reports experiments that were carried out by F. B. in partial fulfillment of the requirements for the degree of a Dr. rer. nat. in the Faculty of Physics and Mathematics of the University of Tübingen.

This work was partially supported by the Deutsche Forschungsgemeinschaft (DFG FOR 1847-A1). Thanks to Anja Horn-Bochtler (LMU München) for contributing the histological sections. We are grateful to Fahad Sultan (Tübingen) and Mitchell Glickstein (London) for their valuable comments on an earlier version of this paper.

